# Risk of HCV infection associated with hospital-based invasive procedures: a systematic review and meta-analysis

**DOI:** 10.1101/2021.11.04.21265936

**Authors:** Paul Henriot, Mathieu Castry, Liem Binh Luong Nguyen, Yusuke Shimakawa, Kévin Jean, Laura Temime

**Author notes:** **Correspondence to:** Paul Henriot, Conservatoire national des arts et métiers (MESuRS Laboratory), 292 rue Saint-Martin, 75003, Paris. **Email address:**.

## Abstract

**Background:** Healthcare settings, where invasive procedures are frequently performed, may play an important role in the transmission dynamics of blood-borne pathogens when compliance with infection control precautions remains suboptimal. This study aims at better understanding and quantifying the role of hospital-based invasive procedures on hepatitis C virus (HCV) transmission.

**Methods:** We conducted a systematic review and meta-analysis to identify recent studies reporting association measures of HCV infection risk that are linked to iatrogenic procedures performed in hospital settings. Based on expert opinion, invasive procedures were categorized into 10 groups for which pooled measures were calculated. Finally, the relationship between pooled measures and the country-level HCV prevalence or the Healthcare Access and Quality (HAQ) index was assessed in meta-regressions.

**Findings:** A total of 71 studies were included in the analysis. The most frequently evaluated procedures were blood transfusion and surgery (60 and 37 studies, respectively). The pooled odds ratio (OR) of HCV infection varied widely, ranging from 1·46 (95%CI: 1·14-1·88) for dental procedures to 3·84 (2·07-7·12) for wound care. The OR for blood transfusion was higher for transfusions performed before 1998 (3·77, 2·42-5·88) than for those without a specified date (2·26, 1·81-2·81). Finally, the country-level overall risk for all procedures was significantly associated with HCV prevalence, but not with the HAQ index. In procedure-specific analyses, the HCV infection risk was significantly negatively associated with the HAQ for endoscopy and positively associated with HCV prevalence for endoscopy and surgery.

**Interpretation:** Various invasive procedures were documented to be significantly associated with HCV infection. Our results provide a ranking of procedures in terms of HCV risk that may be used for prioritization of infection control measures, especially in high HCV prevalence settings.

**Funding:** INSERM-ANRS (France Recherche Nord and Sud Sida-HIV Hépatites)

Research in context

Evidence before this study
We searched PubMed for systematic review or meta-analyses articles published between Jan 1, 2000 and Oct 31, 2021, and focused on iatrogenic risk factors associated with HCV infection in hospitalized patients, using “hepatitis C”, “risk” and “hospital” as keywords. We found a single comprehensive meta-analysis assessing HCV risk factors and reporting pooled risks estimates for hospital-based procedures. However, this meta-analysis was not specific to in-hospital procedures, based on articles published between 1989 and 2013, and did not strictly follow the PRISMA guidelines.

Added value of this study
Our study presents an up-to-date assessment of the risks of HCV infection associated with different hospital-based procedures. Results suggest that iatrogenic HCV infection risk remains important, in studies that were recently conducted. We underline the importance of this risk in high prevalence settings and provide a potential prioritization of procedures associated with HCV infection that may help implementing infection control measures efficiently.

Implications of all the available evidence
This study shows that iatrogenic procedures remain at risk of transmitting blood-borne pathogens, in particular HCV. Despite the availability of highly efficacious direct acting antivirals to treat chronic HCV infection, the prevention of iatrogenic HCV infection, through strictly complying with the established standard precautions, remains essential. These results could be used in modelling studies to help find optimal and efficient control measures to reduce in-hospital HCV transmission.

## Introduction

Hepatitis C virus (HCV) is mainly a blood-borne virus associated with an estimated global sero-prevalence of 2.5%.^**1**^ However, wide between-country discrepancies are observed, with Egypt and Pakistan having high anti-HCV prevalence in the general population.^1^ Chronic HCV infection may lead to serious complications like cirrhosis or hepatocellular carcinoma (HCC), with about 20% developing HCC.^**2**^ Highly effective and well-tolerated direct-acting antiviral treatments (DAAs) are now available, but these treatments remain costly. An estimated 79% of people chronically infected with HCV worldwide still did not know their status in 2019,^3^ limiting the population impact of DAAs. In addition, the residual risk of HCC is suspected to remain relatively high among patients who cured chronic HCV infection.^4^ Moreover, HCV treatment should not be the only way of fighting new infections, as accumulating evidence suggests that even after the DAA therapy-induced sustained virological response (SVR) reinfection can occur, especially in people who inject drugs.^5^ Therefore, prevention measures to limit the risk of HCV contamination remain key in the global response against HCV.^6^

Although injection drug use has been established to be the most important risk factor for acquiring a new HCV infection,^7^ healthcare settings may play an important role in HCV transmission dynamics, due to the high frequency of invasive procedures and over-representation of HCV-infected individuals among hospitalized patients. Medical procedures have been linked to multiple HCV outbreaks worldwide, for instance in Egypt^8^, India^9^ or the United States.^10^ In healthcare settings where compliance with infection control measures is suboptimal, patients may still become infected following blood transfusion or haemodialysis.^11,12^ Previous hospitalization has been identified as a major risk factor for HCV infection in many countries^13^.

In this context, a better control of hospital-acquired infections would significantly help in the fight against HCV epidemics. This requires a clear understanding of the role played by different at-risk procedures within healthcare settings, in order to effectively focus control measures and efforts. Here, we conduct a systematic review and meta-analysis of the current evidence regarding the strength of association between HCV infection and a wide array of hospital-based invasive procedures, in order to propose a prioritization of iatrogenic procedures and better understand their role in HCV transmission.

## Methods

### Search strategy and selection criteria

We searched three online databases (PubMed, Web of Science, Scopus) for studies published between January 2000 and December 2020 using the keywords “hepatitis”, “risk factor”, “hospital”, and “procedure” as main lexical fields (Suppl Table S1). This study was registered in PROSPERO in February 2021 (ID: CRD42021224886), and is reported according to PRISMA guidelines.

Studies were eligible if: (i) Exposure group was composed of adults, either in or out-patients; (ii) they reported measures of association (odds ratios, ORs; risk ratios, RRs; or prevalence ratios, PRs) by comparing the proportion of people positive for HCV RNA or anti-HCV antibody between a group who underwent a specific health-care procedure and another group not exposed to the procedure. There was no restriction in terms of the study design.

Studies were excluded if: (i) They were focused on paediatric patients, drug users, or blood donors only, to avoid bias linked to either age-constraint or repetitive at-risk habits; (ii) patient inclusion started before 2000; (iii) they were not written in French or English; (iv) they did not present original results.

Based on the inclusion and exclusion criteria, articles titles and abstracts were screened by two independent investigators (MC and PH) using the Covidence review tool.^14^ Full-text articles were then retrieved and assessed for eligibility by the same two authors. Any conflict in articles screening or full-text assessment were resolved by a third senior researcher (KJ or LT).

For each study, the following data were extracted by PH: (i) Total number of patients; (ii) patients type; (iii) study design; (iv) measures of association and sample size of the control/exposed groups

### Data analysis

After studies selection, the procedures reported were aggregated into 10 groups based on medical expert opinion (Table 1). Risk of blood borne infection was supposed to be the same within these groups. When two or more procedures classified in the same group were assessed within the same study, a pooled group-level measure was computed using a fixed-effect model since these estimates were calculated based on the same population. Measures associated with the same procedure but reported across different populations (e.g., by gender) were considered independently.

**Table 1:** Aggregation of procedures found within selected articles. Procedure names are reported as mentioned in each of the articles.

| Group | Procedures within group |
| --- | --- |
| *Surgery* | Surgery, operation, bloody operation, splenectomy, laparoscopy, organ biopsy, biopsy, ligation of oesophageal varices, caesarean section |
| *Transplantation* | Transplantation, renal transplantation |
| *Blood transfusion* | dated blood transfusion / non-dated blood transfusion |
| *Intravenous (IV) / Catheter* | Cholangiography, IV pyelography, IV injection, IV-line, sclerotherapy, catheter, central venous catheter, cannula, phlebotomy |
| *Haemodialysis* | Haemodialysis |
| *Wound care* | Wound suture, wound care |
| *Injection* | IM injection, percutaneous injection, subcutaneous injection, injection/vaccination, injection |
| *Other procedures* | EMG, haemorrhoids treatment, tapping ascites, abscess drainage, abortion |
| *Endoscopy* | Endoscopy, colonoscopy, gastroscopy |
| *Dental care* | Tooth extraction, dental anaesthesia, dental procedure, dental care, tooth filling |

We performed a meta-analysis to compute pooled OR estimates of the risk of HCV infection associated with each procedure group using the R “meta” package.^15^ Procedure-specific forest plots were visually inspected for potential outliers. Measures that were not ORs (RRs and PRs) were considered to be equivalent to ORs and included in the same analyses. When both adjusted ORs (AORs) and crude ORs (CORs) were available in the same study, the AORs were preferred. In a same study, if risk estimates were available for dated and undated (without specified date) blood transfusion, estimates associated with undated blood transfusion were preferred.

In a second step, pooled ORs for low HCV prevalence (< 5%) and high HCV prevalence (> 5%) countries were compared through subgroups analysis for each procedure group. Prevalence data for each country were collected from the MapCrowd online global data on hepatitis C (Suppl Table S7).^16^ Pooled ORs were stratified by country for the 2 most represented procedure groups. In addition, subgroups analyses were performed to compare estimates between cohort and non-cohort studies.

Thirdly, the pooled OR for blood transfusion based on transfusions performed with either recent dated or an unspecified date of realization was compared to the one based on transfusions performed before 1998, which is the latest cut-off date used within selected articles. We assumed cut-off dates found in the studies selected to reflect local implementation of mandatory HCV screening in blood donors.

Finally, meta-regressions were performed to investigate the potential effect modifier of: (i) the HCV prevalence level, and (ii) the Healthcare access and quality (HAQ) index, a 0 to 100 score quantifying the strength of healthcare quality and access based on amenable mortality data in each country (Suppl Table S7).^**17**^

Pooled estimates were computed using random effect models considering our aim to generalize results beyond the selected studies. Each of these estimates was calculated together with the heterogeneity level (I^2^ statistics). Comparisons were performed using Cochran’s Q test.^18^

The risk of bias was assessed by PH on the articles included. Nine bias assessment criteria were adapted from Lam *et al*. (2016c)^19^, and publication bias was assessed by testing for asymmetry in the overall and procedure-specific funnel plots.^19^

## Results

Among the 1,961 initially identified studies, 71 were included in the review and meta-analysis^21-91^, as described in the flow diagram (Fig. 1). The total number of participants across all selected studies was 120,734.

**Figure 1:**
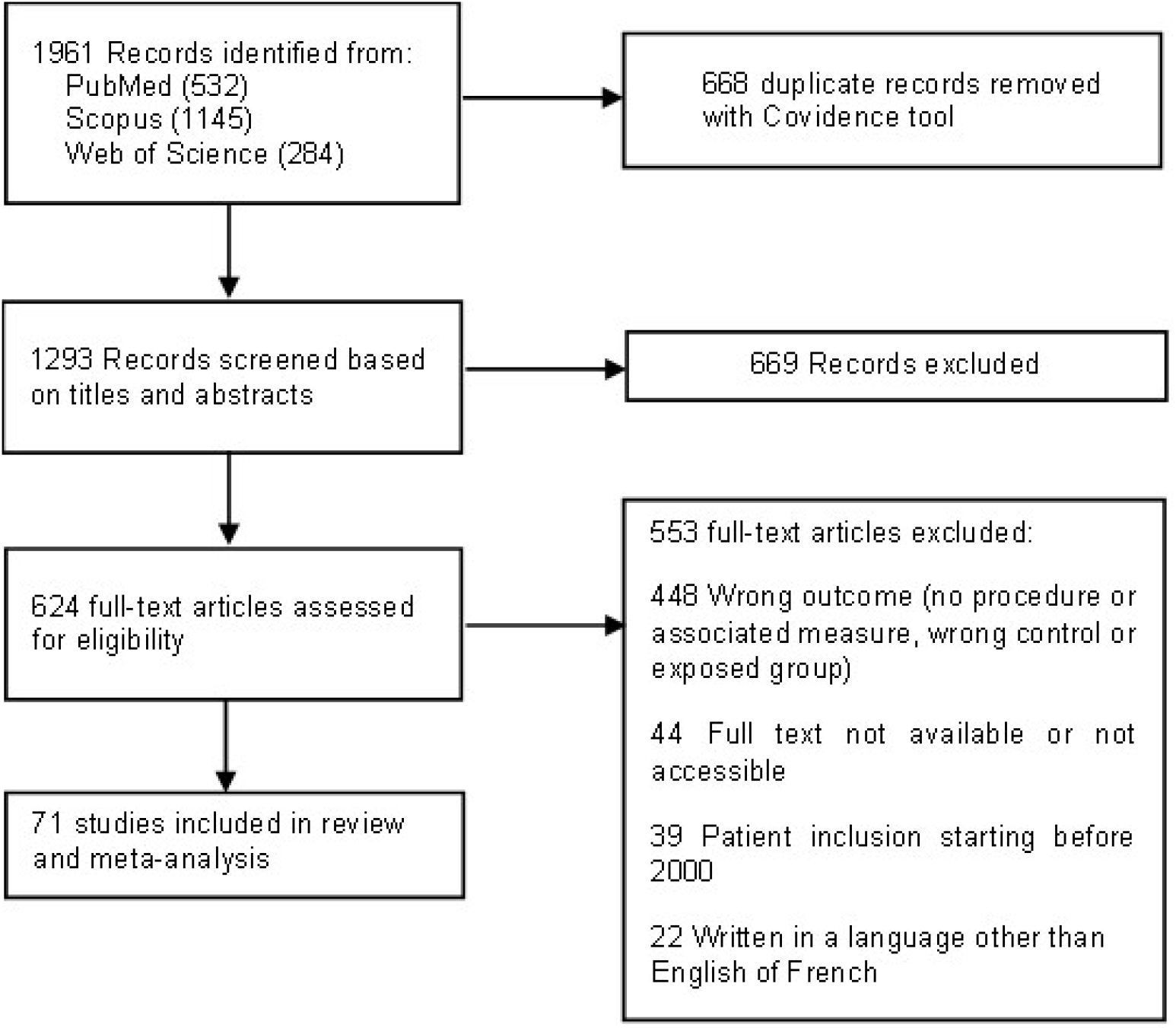
Study selection.

Fig. 2.a shows an increase in publications related to HCV risk assessment within hospitals between 2000 and 2015, while the number of publications remained stable between 2015 and 2020. Together, Pakistan (fourteen studies) and Egypt (nine studies) represented one third of all included studies (Fig2.c). Over a fifth of studies were focused on haemodialysis patients (21·9%) (Fig. 2.b). Most studies were cross-sectional (55%) or case-control (22·5%) studies (Fig. 2d).

**Figure 2:**
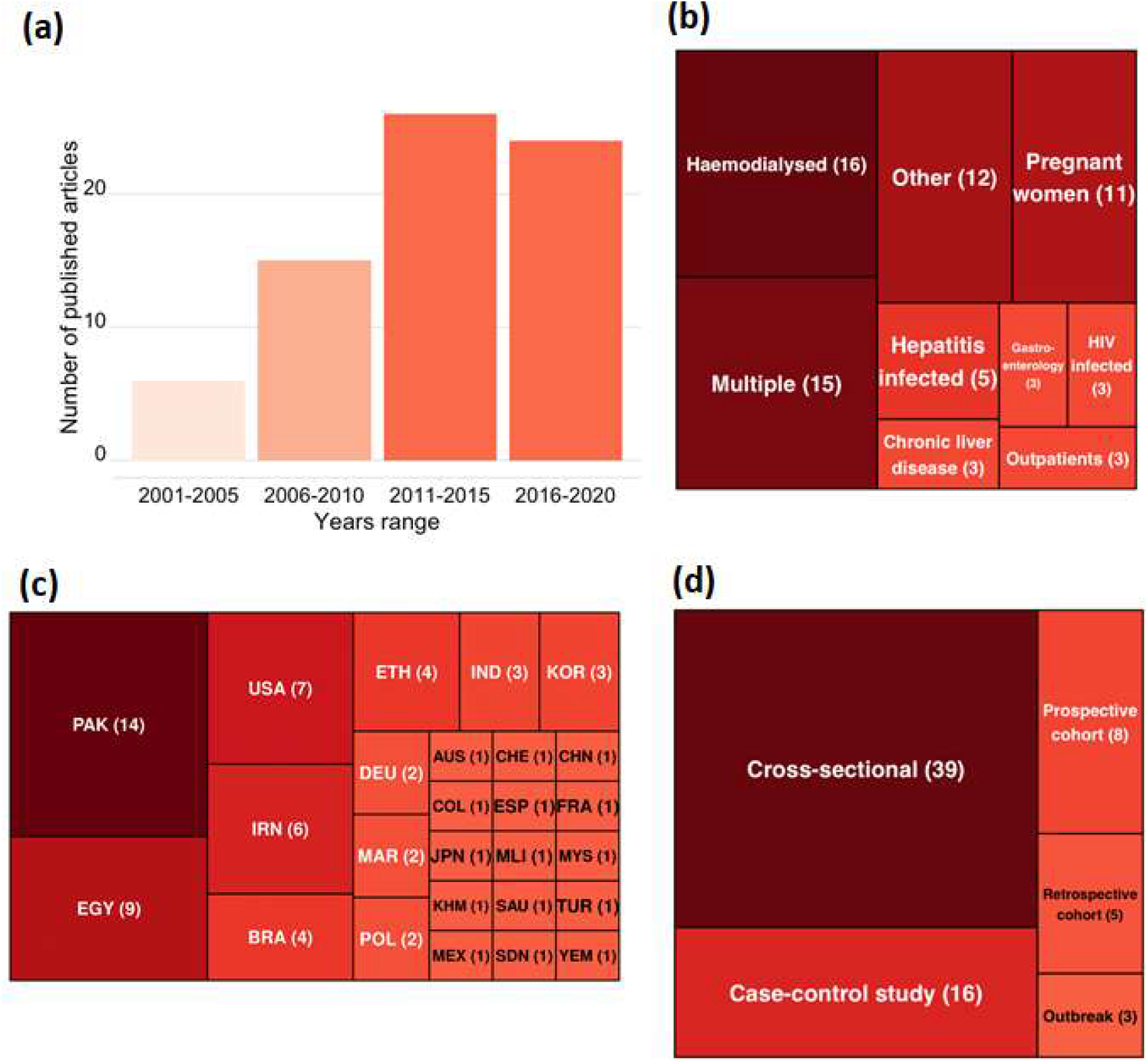
Study characteristics.. Number of included studies by: (a) year of publication; (b) type of patients; (c) country; (d) Study designs.

Forty-five different procedures were assessed (Table 1). The number of observations per each of the ten procedure groups ranged from five to sixty, with surgery and transfusion as the largest groups (respectively 37 and 60) (Fig. 3, Suppl Table S6). Visual inspection of procedure-specific forest plots (Suppl Fig. S6 to S15) led to the exclusion of a single outlier for haemodialysis (Suppl Fig. S6).

**Figure 3:**
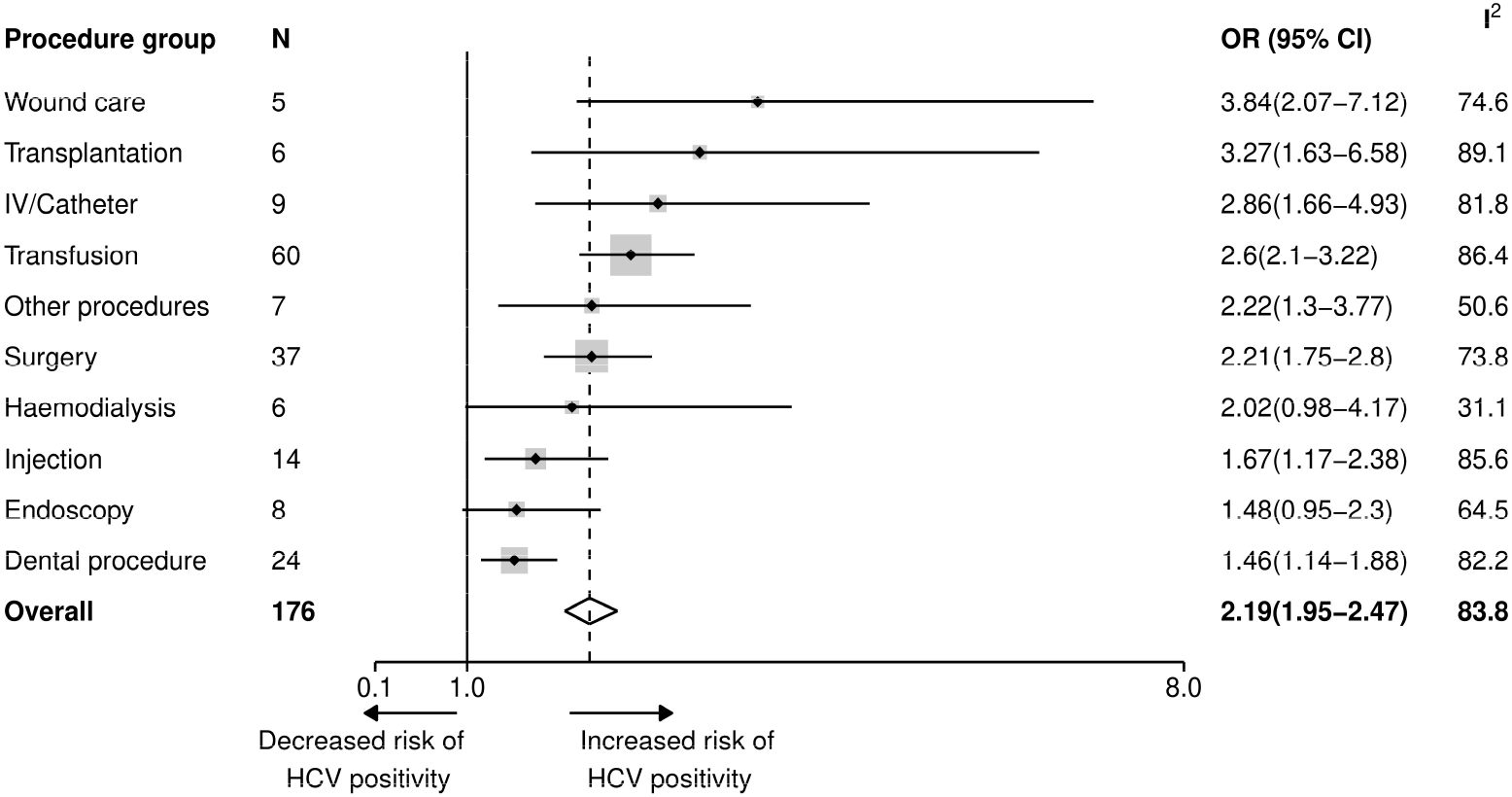
Forest plot reporting pooled OR estimates for HCV infection risk associated with different groups of iatrogenic procedures. Points correspond to average estimates and error bars correspond to 95% CI of these estimates. The solid line represents the limit for which OR = 1 and the dotted line the value of the overall estimate. The total number of observations used for each pooled OR calculation is specified in the column *N* and represented as the size of the grey area around each point estimate; the heterogeneity for each estimation is depicted in the column *I*^*2*^. A given study could be used in the OR estimation of multiple procedures groups. Procedures are sorted based on the value of their associated mean estimate, from the highest to the lowest.

All pooled ORs were significantly superior to one, except for endoscopy and haemodialysis.

Procedure-specific analyses showed that the risk of HCV infection differed significantly according to procedure groups (Q = 22·48, p < 0·01), with pooled OR estimates ranging from 1·46 (CI 95% [1·14-1·88]) for dental procedure up to 3·84 (CI 95% [2·07-7·12]) for wound care. The IV/Catheter, wound care and transplantation groups had OR estimates ranging from 2·86 (CI 95% [1·66-4·93] for IV/Catheter) to 3·84 (CI 95% [2·07-7·12] for wound care), representing moderate/high risk groups. The haemodialysis, surgery, other procedure, and transfusion groups had intermediate pooled ORs, with estimates of 2·02 (CI 95% [0·98-4·17]), 2·21 (CI 95% [1·75-2·8]), 2·22 (CI 95% [1·3-3·77]),2·6 (CI 95% [2·1-3·22]) respectively. Injection was associated with a low/moderate risk of HCV infection with an estimate of 1·69 (CI 95% [1·19-2·41]). Finally, the dental procedure and endoscopy groups presented the lowest risks of getting HCV infected (1·46 CI 95% [1·14-1·88] and 1·48 CI 95% [0·95-2·30] respectively).

Country-stratified analyses were performed for transfusion and surgery (Fig. 4). For these two procedure groups, estimated ORs varied widely between countries. The highest risk of HCV infection by blood transfusion was reported in Germany (OR = 5·39 CI 95% [2·67-10·89]) but this estimate corresponds to only one study and is associated with transfusions performed before 1991. Other countries associated with relatively high HCV infection average risk for blood transfusion were Egypt (OR = 5·16 CI 95% [1·86-14·28]), Iran (5·11 [1·04-24·99]), Turkey (4·50 [1·71-11·86]) and Malaysia (4 [2·18-7·35]). Nevertheless, associated confidence intervals were quite wide. There were less high-risk countries concerning surgery, for which Egypt and India were associated with the highest risk (4·27 [2.03-8·98] and 4·62 [0·82-26·02]).

**Figure 4:**
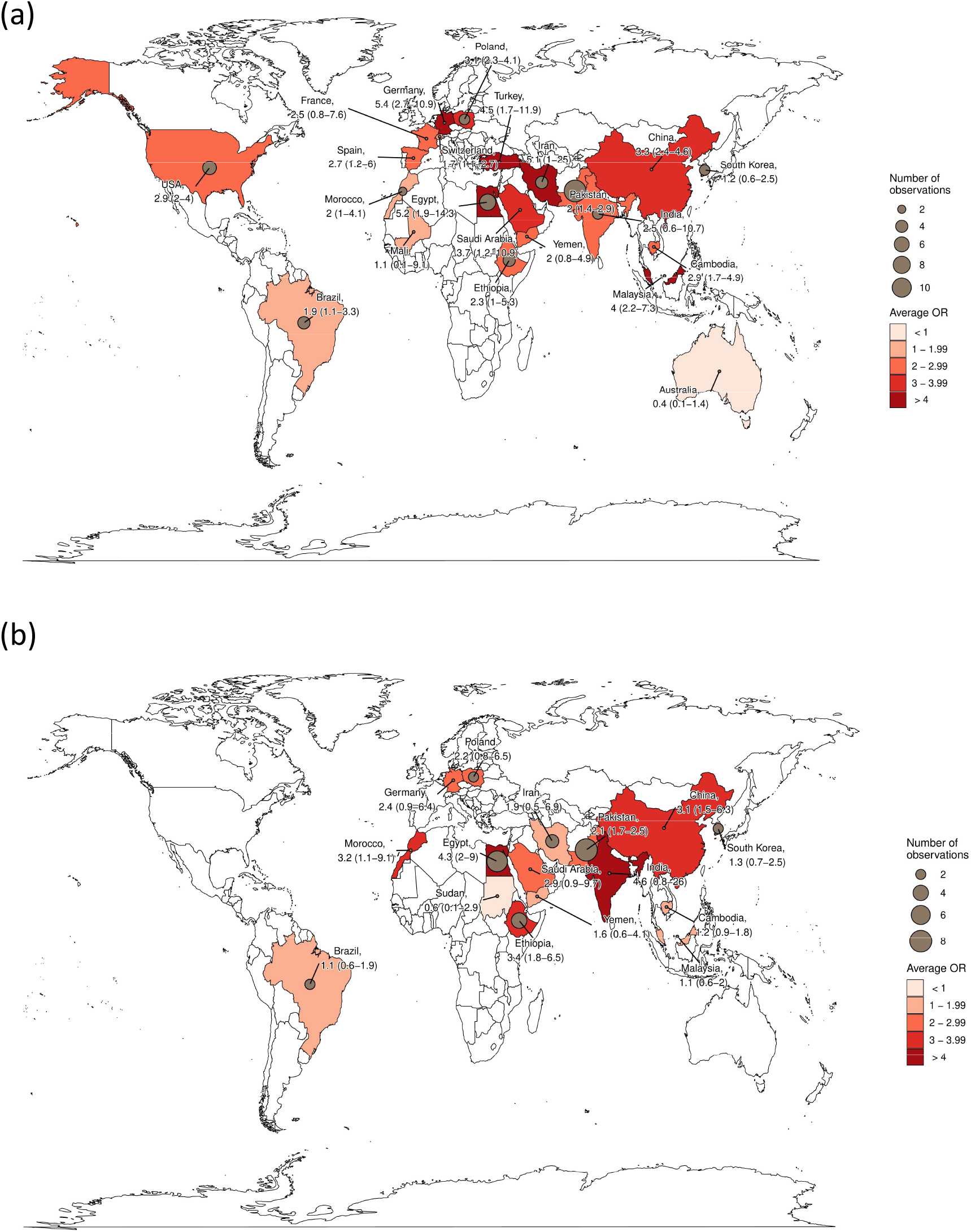
Maps of risks of HCV infection for: (a) Blood transfusion (b) Surgery in various countries. Brown points correspond to the number of observations used to compute a given pooled OR. Each country is coloured based on the value of the estimated OR following a light red to dark red colour gradient. Values of average estimates are reported as well as their 95% CI.

Overall, average estimates were always higher in high prevalence countries (Fig. 5) though this difference was not significant (Q=2·11, p=0·15), Nevertheless, a significant difference was observed for endoscopy (Q=9·40, p-value=0·002), surgery (Q=4·54, p-value=0·03) and injection (Q=5·40, p =0·02). These results were supported by procedure-specific meta-regressions (Suppl Table S3) showing prevalence to have a significant positive effect on the OR level of the endoscopy (p < 0·01) and surgery (p=0·011) groups. Prevalence was also found to have a significant positive effect on the risk of iatrogenic HCV infection as a whole (p < 0·01, Suppl Table S5). On the contrary, no overall association was found between HAQ level and risk of iatrogenic infection, while in procedure-specific analyses a negative significant impact of the HAQ level was observed for endoscopy only (p=0·02, Suppl Table S4).

**Figure 5:**
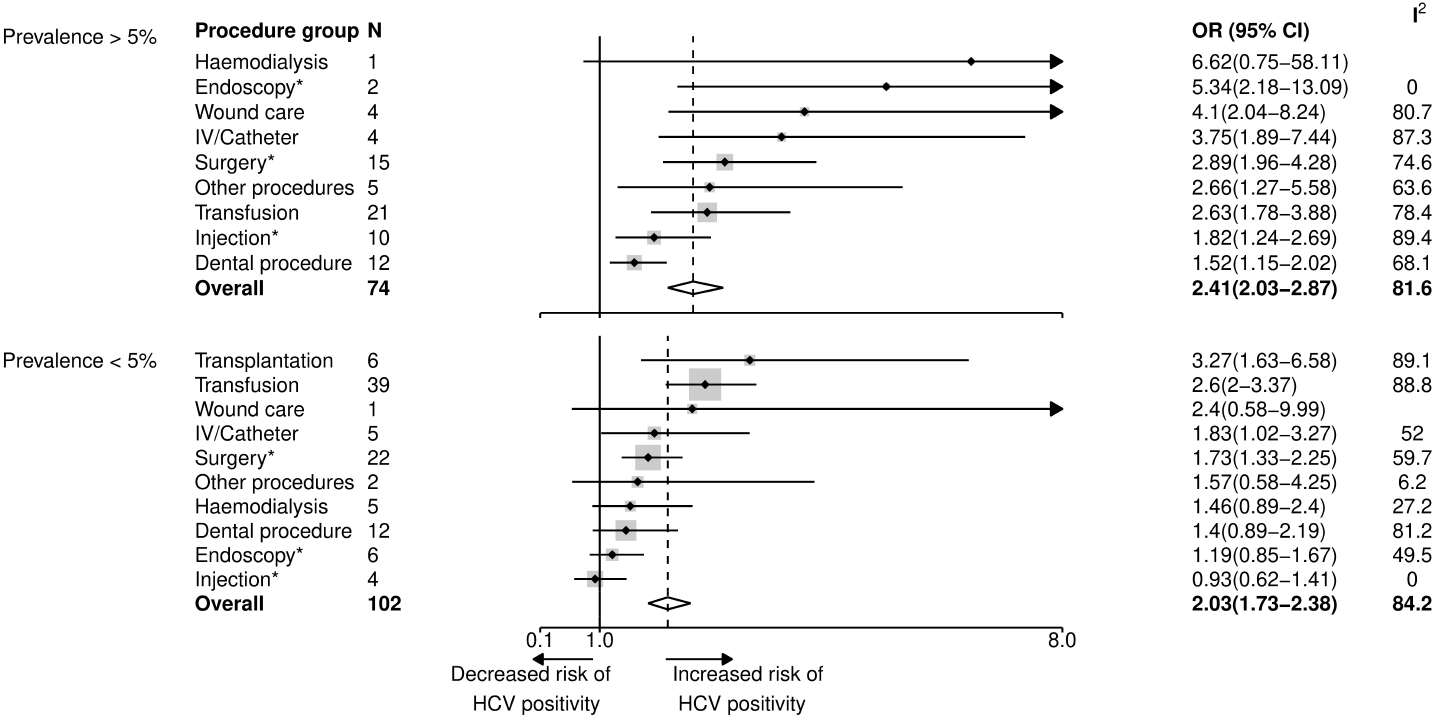
Forest plot reporting pooled OR estimates for HCV infection risk associated with different groups of iatrogenic procedures for high HCV prevalence countries (>5%) vs. low HCV prevalence countries (<5%). The high prevalence group includes Egypt, Pakistan and Mali. Points correspond to average estimates and error bars correspond to 95% CI of these estimates. The solid line represents the limit for which OR = 1 and the dotted line the average value of the overall estimate. The total number of observations used for each pooled OR calculation is specified in the column *N* and the heterogeneity for each estimation is depicted in the column *I*^*2*^. A given study could be used in the OR estimation of multiple procedures groups. Procedures are sorted based on the value of their associated mean estimate, from the highest to the lowest. There was no measure reported for transplantation in high HCV prevalence countries. *: procedures for which a significant different was found between high and low prevalence countries; Blanks within the *I*^*2*^ column correspond to absence of heterogeneity because of procedure group counting only one observation.

For blood transfusions performed before 1998, 17 studies were selected for the pooled OR calculation, whereas there were 48 studies available for undated blood transfusions (Suppl Fig. S3 and S4). The estimated pooled OR of HCV infection associated with blood transfusions performed before 1998 (OR = 3·77 CI 95% [2·42-5·88]) was significantly higher than the pooled OR associated with undated blood transfusions (2·28 [1·82-2·84], p = 0·05, Q =3·97)

Across all studies, bias was low for most criteria, but some studies presented a potentially high risk of bias for criteria related to exposure assessment and to potential confounding (Suppl Fig.S1 and Suppl Table S2). No single study had high bias risk for all criteria. Nevertheless, the overall funnel plot showed asymmetry and the associated Egger test was highly significant, suggesting potential publication bias (Suppl Fig. S2).

Average ORs in non-cohort studies were found to be higher than in cohort studies (2·26 CI 95% [1·98-2·57] vs. 1·92 [1·47-2·50]) but this difference was not significant (Q = 1·16, p = 0·28). Per-procedure comparisons between cohort and non-cohort studies only showed a significant difference for the transplantation (Q = 6·32, p = 0·01) and the injection (Q = 4·62, p = 0·03) group, for which cohort studies was found to be associated with lower estimates.

## Discussion

In this systematic review and meta-analysis, we identified 71 studies assessing the association between hospital-based invasive procedures and the risk of HCV infection, representing a total of 120,734 participants. The overall strength of evidence remained of limited confidence as the bias analysis found a lack of high-quality studies. Nevertheless, we estimated pooled ORs of HCV infection that were significantly associated with most invasive procedures performed in hospitals. Our results suggest a prioritization of iatrogenic procedures: wound care was associated with the highest risk of HCV infection whereas dental procedures and endoscopy were associated with the lowest risk. We also underlined a large between-procedure and between-country variability, and showed that the per-procedure risk tended to be higher in countries with high HCV prevalence, while the level of healthcare quality and access in the country (as measured by the HAQ) only appeared to play a minor role.

The geographical coverage of selected studies was in line with global observed prevalence levels, with a third of these studies coming from the two countries with the highest HCV prevalence worldwide (Egypt and Pakistan). In addition, the most represented population was composed of haemodialyzed patients, for which the risk of HCV infection is historically susbtantial.^92^ Regrettably, only few studies used cohort data.

The estimated per-procedure risks seemed to be mostly in line with the available literature concerning HCV. Generally, the risk was higher for procedures with frequent blood-contact.

Surprisingly, we found that the highest risk of infection was found for wound care. The estimate of the HCV infection risk through sutures found in the only previous meta-analysis by El Ghitany et al. was lower than our estimate, but increased in the most recent studies.^13^

The risk associated with organ transplantation was found to be quite high, considering that most of countries now require HCV testing in donors.^92^ This might result from either lack of information on the procedure realization date or the type of study design used to assess this risk, as it was found to be much lower considering only cohort studies. Similarly, the intermediate risk we found for blood transfusion could be explained by the fact that there is still high discrepancy in systematic screening of blood products between countries (only 80% of donations are screened in low-income countries).^93^ Haemodialysis was also associated with an intermediate risk of HCV infection. Our estimate was within the same order of magnitude than the one reported in the previous meta-analysis.^13^

We found injection to be one of the lowest at-risk procedures. Iatrogenic injection was described in the past as associated with a high risk of HCV infection, in particular because of the reuse of contaminated syringes,^8^ but this risk might have dropped over the last decade, in particular after the publication of multiple WHO and CDC guidelines for safe injections.^94^

Finally, dental procedures and endoscopy were found to be the lowest risk groups, in line with current literature findings. Indeed, only a few cases of HCV contamination after endoscopy have been described and dental practices are often at low risk of contamination.^92,95^ This result is also consistent with previous estimations showing low risk associated with these two procedures.^13^

Country-level prevalence was overall found to be related to a higher risk of HCV contamination, although it was only significantly associated with endoscopy and surgery risks when looking at specific procedures. The lack of significance for other procedures may be explained in several ways. First, many countries were represented by less than 3 studies, leading to a lack of power for some procedures and countries and less accurate OR estimates. Second, these non-significant associations may also indicate the presence of other causes of heterogeneity. In particular, there may be high differences in terms of prevalence between hospital settings within the same country. Observed between-country variations may also result from different infection control practices; we explored this assumption using the HAQ index, which is internationally validated and available for all countries in our analysis. However, we found no significant relationship between the risk of getting HCV contaminated and this index, except for the risk linked to endoscopy. First, the HAQ index may not be the right indicator to accurately reflect compliance with infection control measures within hospitals. Second, there could be a high heterogeneity in this compliance between settings within the same country, that this index is unable to capture.

Our study carries several methodological as well as more study-related limitations.

First, no grey literature or papers in languages other than French or English were included in this review, resulting in a potential information loss. Some other methodological simplifications could have influenced our final results. In fact, pooling adjusted and non-adjusted measures could have led to overestimation of risks associated with some procedures. Our results also highly depend on the initial procedures classification we proposed, which may contribute to the high between-study heterogeneity we observed for many procedure groups. However, this classification was necessary due to the overall high number of unique procedures identified in the review. Moreover, the high heterogeneity could also result from other sources (e.g., prevalence or infection control).

This study highlighted several caveats in the existing literature. Bias analysis showed a lack of high-quality studies. On the one hand, 90% of studies presented at least a probably high bias concerning the assessment of exposure to hospital-based procedures, mostly using questionnaires to collect risk factors for HCV infection. This is consistent with the distribution of studies design, since almost 80% of studies were either cross-sectional or case-control studies. This over-representation of non-longitudinal designs may in particular have led to overestimated ORs of HCV infection associated with invasive iatrogenic procedures, due to differential recall bias between cases and controls. We investigated this through a separate assessment of pooled ORs in non-cohort and cohort studies, lower ORs were indeed found in cohorts for injection and transplantation. However, ORs for all other procedures, as well as overall ORs, did not differ significantly between cohorts and non-cohorts, supporting our choice to consider all study designs together. On the other hand, more than 50% of studies included a potential risk of bias due to incomplete or missing use of adjustment variables when estimating measures of association between HCV infection and iatrogenic procedures. Overall, heterogeneity in measures associated with iatrogenic procedures was shown and may reveal potential publication bias. In particular, non-significant risk measures might have been voluntarily neglected resulting, again, in an overestimation of the risk associated with these two groups of procedures.

For some procedure groups, only few data were available, which led to important uncertainty in the associated OR estimates. Half of procedure groups had less than ten risk measures, wound care being the procedure for which this number was the lowest with only five studies. Also, some procedures like surgery might have described operations during which multiple systematic procedures were performed and not taken into account (pre-surgical anaesthesia, …). More generally, we rarely had access to measures of risks associated with a unique realization of each procedure. Patients might have undergone the same procedure multiple times but this information was not taken into account. In particular, the risk of getting HCV infected during haemodialysis is highly related to the duration of haemodialysis and the risk related to blood transfusion is somehow positively proportional to the number of received transfusions.^96^ These limitations may have caused overestimation of the risk associated with hospital-based procedures.

In addition, the date of realization of a procedure was not always available, and some measures might have been based on procedures performed a long time ago, thus leading again to an overestimation of the risk. Indeed, we found the pooled OR of blood transfusions performed before 1998 to be higher than the pooled OR of undated blood transfusions, strongly suggesting that the risk was higher for older procedures - all the more so that the undated procedures could also have included old procedures.

In conclusion, despite the uncertainty in our estimates and the probable decrease in the general risk of HCV infection through hospital-based procedures over the last twenty years, this work shows that healthcare settings remain an important gateway of HCV infection and underline the importance of implementing efficient infection control.

Our results suggest a risk-based ranking for iatrogenic procedures, wound care being the most at-risk procedure, and confirm the important role of blood screening in decreasing the risk of HCV infection. We also show that the strength of association between HCV infection and iatrogenic procedures tend to be higher in high prevalence countries. This could partly be explained by the high global heterogeneity in infection and prevention control knowledge in healthcare workers, especially concerning bloodborne pathogens. ^97^

Two main tracks appear to be key to control HCV iatrogenic infections. First, knowledge of existing infection control measures should be improved in order to make them fully effective. Second, prevention efforts should be specifically targeted at high-risk procedures, such as wound care, transplantation or procedures involving intravenous injection, and in high HCV prevalence settings. In this regard, our work provides support to identify the procedures on which these efforts should be focused. Our results may also prove useful for future modelling studies assessing the effectiveness of control measures to limit HCV transmission in healthcare settings.

In 2016, the World Health Organization (WHO) set targets for global HCV elimination as a public health problem by 2030 – an 80% reduction in incidence and a 65% reduction in mortality from 2015 levels.^98^ In order to reach these targets, high-prevalence countries such as Egypt and Pakistan have engaged in elimination programmes based on large-scale test-and-treat campaigns.^99^ By helping efficiently scale up prevention interventions, our work has the potential to improve the cost-effectiveness of these campaigns, reaching the WHO targets faster and more easily.

## Data Availability

All data used in the present work is available in the supplementary material.

## Contributors

PH, KJ and LT conceived the study, wrote the protocol and data analysis plan. PH and MC screened the articles and conflicts were resolved by LT and KJ. Clinical input and HCV expertise were provided by LBLN and YS. PH did the statistical analysis. PH, LT and KJ interpreted the data. PH wrote the first draft of the report with input from LT, KJ, MC, LBLN and YS. All authors had full access to all the data in the study and had final responsibility for the decision to submit for publication. PH, KJ and LT accessed and verified the data.

## Data sharing

Data summary will be available in the supplementary appendix. The statistical analysis plan will be available with publication upon reasonable request to the corresponding author.

## Acknowledgements

This study was funded by INSERM-ANRS (France Recherche Nord and Sud Sida-HIV Hépatites), grant number 12320 B115.

