## Supplementary appendix for "Risk of HCV infection associated with hospital-based invasive procedures: a systematic review and meta-analysis"

**Remarks on the study protocol registered in Prospero**

The study protocol (Prospero ID: CRD42021224886) was modified on the 21th of June 2021 to clarify inclusion criteria as followed:

*Old version (02/04/2021)*

"Eligible studies have to fulfil the following criteria:
1. Focus on hospitalized in/out-patients
2. Assess risk factors for HCV infection among these patients, and include a quantified measure: odds ratios (ORs) or risk ratios (RRs) or prevalence ratios (PRs)
3. Assess the HCV risk associated with at least one medical or surgical procedure (e.g. injection, sutures, transfusion…)"

*New version (06/21/2021)*

"Eligible studies have to fulfil the following criteria:
1. Focus on hospitalized in/out-patients
2.Assess the HCV risk associated with at least one medical or surgical procedure (e.g. injection, sutures, transfusion…)
3. Assess risk factors for HCV infection among these patients, and include a quantified measure of association between incident or prevalent HCV infection and one or more medical or surgical procedures as compared to a control group unexposed to these procedures: odds ratios (ORs) or risk ratios (RRs) or prevalence ratios (PRs)"

In addition, risk of bias was not assessed by the Newcastle-Ottawa Scale as mentioned in Prospero but was evaluated as proposed by Lam *et al.* (2016c)[1].

| Authors | Exposed + (n/N) | Exposed – (n/N) | OR | 95%–CI | Weight |
|---|---|---|---|---|---|
| De Weggheleire, A. et al. | 36/301 | 121/2699 | 2.91 | [1.74; 4.87] | 3.0% |
| Kundu, A.et al. | 0/9 | 5/41 | 0.80 | [0.69; 0.93] | 3.6% |
| Balew, M. et al. | 2/5 | 35/390 | 5.61 | [0.94; 33.44] | 1.1% |
| Shahriari–Fard, F.et al. | 68/493 | 47/521 | 1.61 | [1.09; 2.38] | 3.3% |
| Amjad, U. et al. | 31/40 | 31/56 | 2.78 | [1.12; 6.91] | 2.3% |
| Aslam, A.et al. | 31/290 | 11/290 | 3.04 | [1.42; 6.52] | 2.6% |
| Khan, U. R. et al. | 36/100 | 83/257 | 1.17 | [0.72; 1.91] | 3.1% |
| Kandeel, A. M. et al. | 7/86 | 2/287 | 12.60 | [2.38; 66.75] | 1.2% |
| Vidales–Braz, B.M. et al. | 9/11 | 37/47 | 1.16 | [0.66; 2.04] | 3.0% |
| Qureshi, H. et al. | 42/723 | 15/723 | 2.90 | [1.51; 5.55] | 2.8% |
| Bouare, B.et al. | 1/13 | 15/217 | 1.12 | [0.14; 9.09] | 0.9% |
| Asemahagn, M. A. | 3/18 | 36/404 | 2.00 | [0.56; 7.21] | 1.6% |
| Murad, E.A.et al. | 14/34 | 70/362 | 2.00 | [0.82; 4.90] | 2.3% |
| da Silva, N. M. et al. | 36/37 | 114/122 | 1.93 | [0.32; 11.60] | 1.1% |
| Jamil, Z. et al. | 156/525 | 41/149 | 1.11 | [0.74; 1.67] | 3.2% |
| Asad, M. et al. | 5/112 | NA/233 | 2.08 | [0.61; 7.06] | 1.7% |
| Jaffery, T. et al. | 3/31 | 2/62 | 3.85 | [0.28; 52.48] | 0.6% |
| Khalid, G.G., et al. | 203/1847 | 305/3145 | 1.00 | [0.82; 1.22] | 3.5% |
| Silva, J. L. et al. | 43/69 | 101/161 | 2.08 | [1.16; 3.73] | 2.9% |
| Paez Jimenez, A .et al. | 4/100 | 1/679 | 34.90 | [3.27; 372.07] | 0.7% |
| Neumeister, A. S. et al. | 8/21 | 38/222 | 2.70 | [1.04; 6.99] | 2.2% |
| Galperim, B. et al. | 60/68 | 187/257 | 3.90 | [1.47; 10.32] | 2.1% |
| Mohsen, A. et al. | 4/111 | 1/111 | 4.00 | [0.42; 37.84] | 0.8% |
| Prasad, M. et al. | 13/131 | 10/251 | 3.30 | [0.95; 11.49] | 1.7% |
| Senosy, S. A., et al. | 21/37 | 35/343 | 2.32 | [1.48; 3.64] | 3.2% |
| Samimi–Rad, K. et al. | 6/7 | 121/153 | 1.59 | [0.18; 13.87] | 0.8% |
| Ba–Essa, E. M. et al. | 9/20 | 176/1037 | 3.67 | [1.23; 10.95] | 1.9% |
| Samimi–Rad, K. et al. | 10/11 | 122/193 | 5.80 | [0.71; 47.22] | 0.9% |
| Akhtar, A.M. et al. | 1/9 | 12/74 | 0.65 | [0.07; 5.68] | 0.8% |
| Amin Elzorkany, K. M., et al. | 51/61 | 415/590 | 1.01 | [0.47; 2.20] | 2.5% |
| Ghias, M., et al. | 89/281 | 24/119 | 9.20 | [2.03; 41.80] | 1.4% |
| Taye, M. et al. | 3/23 | 22/422 | 1.84 | [0.41; 8.30] | 1.4% |
| Mostafa, A.et al. | 7/22 | 28/2155 | 29.10 | [9.59; 88.30] | 1.9% |
| Ver Hoeve, E. et al. | 95/145 | 205/455 | 2.40 | [1.20; 4.78] | 2.7% |
| Russmann, S., et al. | 38/135 | 945/4592 | 1.70 | [1.07; 2.71] | 3.1% |
| Loras, C., et al. | NA/37 | NA/2039 | 2.70 | [1.22; 5.99] | 2.5% |
| Doblali, T.et al. | 22/26 | 78/115 | 2.79 | [1.16; 6.73] | 2.3% |
| Weisbord, J. S. et al. | 4/32 | 41/655 | 2.02 | [0.71; 5.73] | 2.0% |
| Ejeta, E. et al. | 1/34 | 7/387 | 1.40 | [0.10; 19.12] | 0.6% |
| Alavian, S. M.et al. | 96/108 | 517/642 | 1.90 | [1.05; 3.44] | 2.9% |
| Mittal, G. et al. | 26/36 | 42/82 | 1.90 | [1.02; 3.53] | 2.8% |
| AbdulQawi, K., et al. | 12/83 | 45/1141 | 2.69 | [1.20; 6.01] | 2.5% |
| Sohn, H. S. et al. | 64/234 | 78/234 | 0.76 | [0.51; 1.13] | 3.3% |
| Haider, J.et al. | 5/46 | 30/1494 | 5.91 | [2.18; 16.02] | 2.1% |
| Bibi, S.et al. | 102/146 | 106/197 | 1.99 | [1.26; 3.13] | 3.2% |
| Patil, S. R., et al. | NA/116 | NA/25077 | 10.13 | [5.99; 17.12] | 3.0% |
| Lioussfi, Z. et al. | 32/40 | 18/27 | 0.87 | [0.13; 6.02] | 1.0% |
| | | | **2.28** | **[1.82; 2.84]** | **100.0%** |

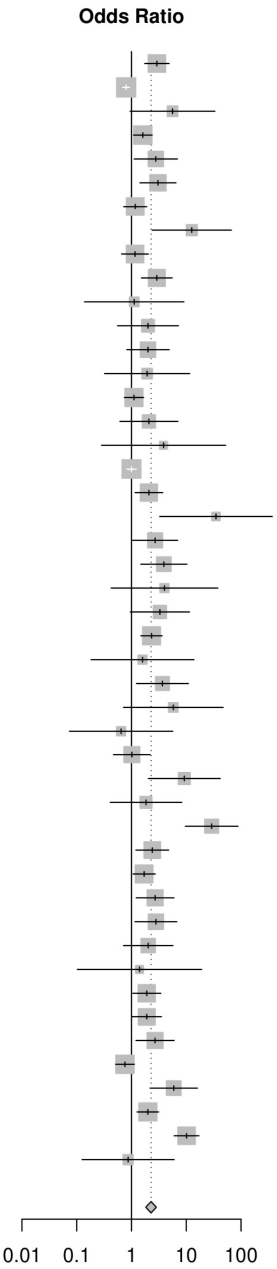

***Figure S1*: Forest plot for undated blood transfusion**

The second and third column report the number of positive and negative procedure-exposed patients (n) over the total number of positive and negative patients (N). NA : Information was not available.

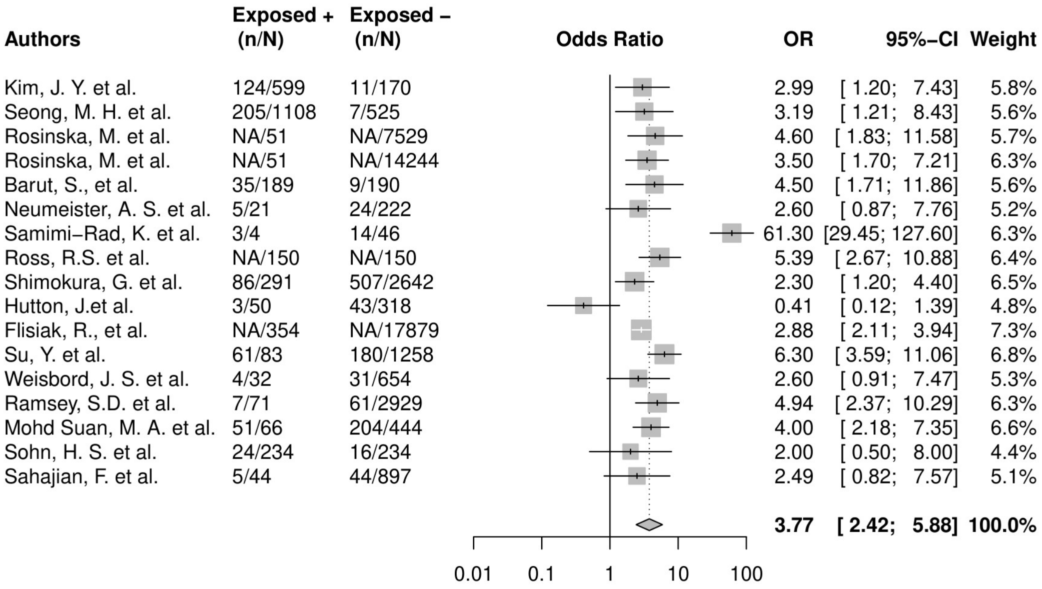

***Figure S2*****: Forest plot for old blood transfusion (before 1998)**

The second and third column report the number of positive and negative procedure-exposed patients (n) over the total number of positive and negative patients (N). NA : Information was not available.

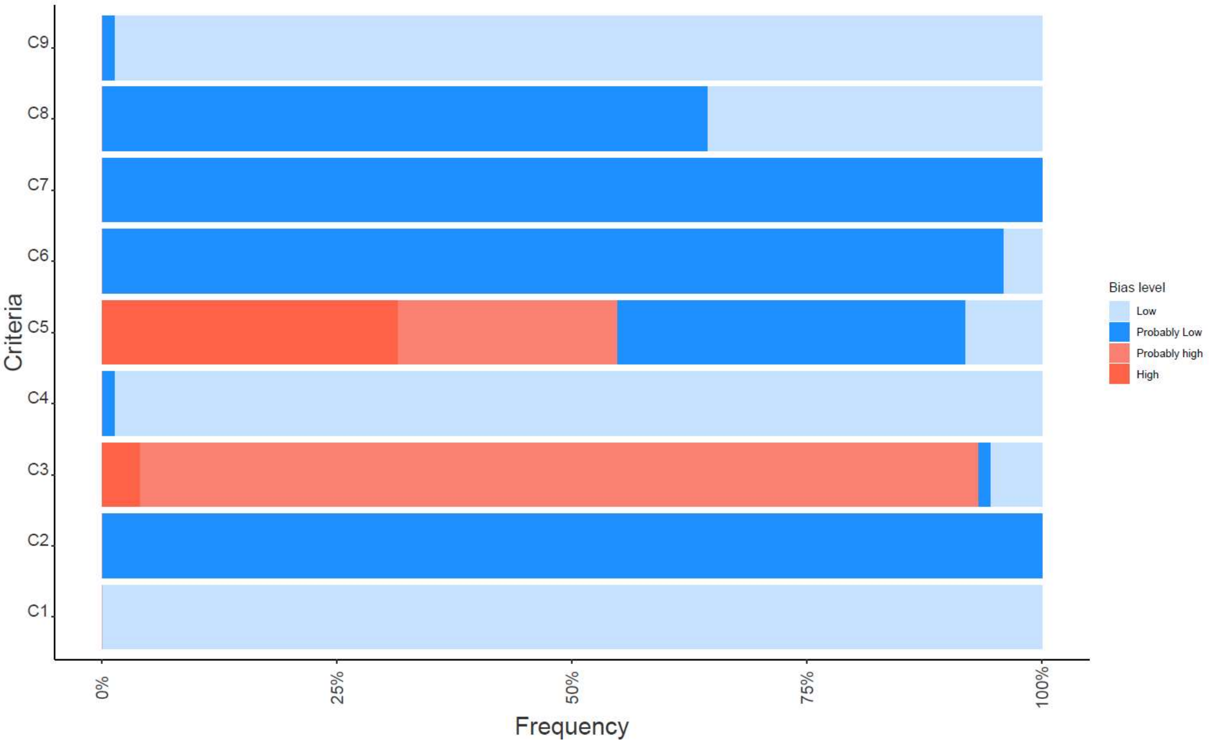

***Figure S3:*** **Frequency of the four bias levels for each considered criterion**.

**C1**: *Are the study groups at risk of not representing their source populations in a manner that might introduce selection bias?* **C2**: *Was knowledge of the group assignments inadequately prevented (i.e., blinded or masked) during the study, potentially leading to subjective measurement of either exposure or outcome?* **C3**: *Were exposure assessment methods lacking accuracy?* **C4**: *Were outcome assessment methods lacking accuracy (i.e., HCV status obtained by anti-HCV or HCV RNA testing)?* **C5**: *Was potential confounding inadequately incorporated (i.e., were outcomes adjusted on other factors)?* **C6**: *Were incomplete outcome data inadequately addressed?* **C7**: *Does the study report appear to have* *selective outcome reporting?* **C8**: *Did the study receive any support from a company, study author, or other entity having a financial interest in any of the exposures studied?* **C9**: *Did the study appear to have other problems that could put it at a risk of bias?*

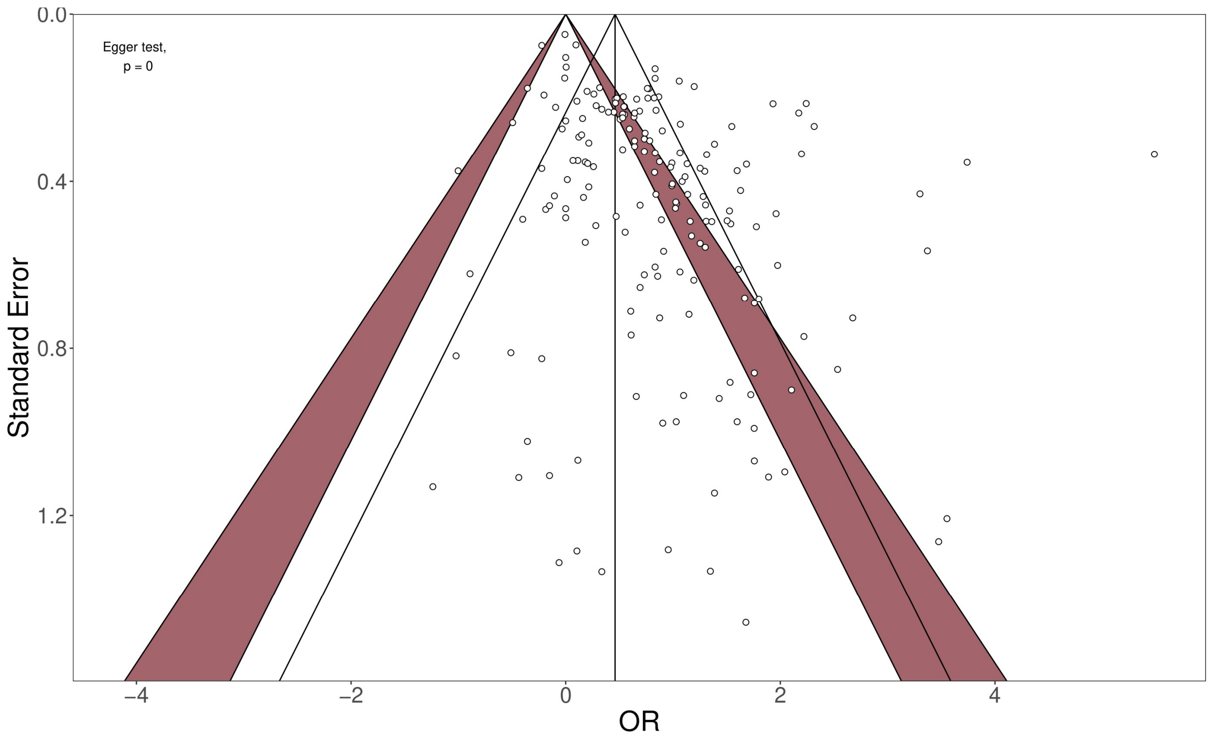

***Figure S4*: Contour-enhanced funnel plot for all measures within selected studies**
These plots show points of individual odds ratios collected in selected studies against associated standard errors. Red-coloured region corresponds to area of statistical significance (p<0.05) for the represented measures. The solid vertical line represents the overall effect associated with the random-effect model, and the solid triangle represents the associated 95% CI.

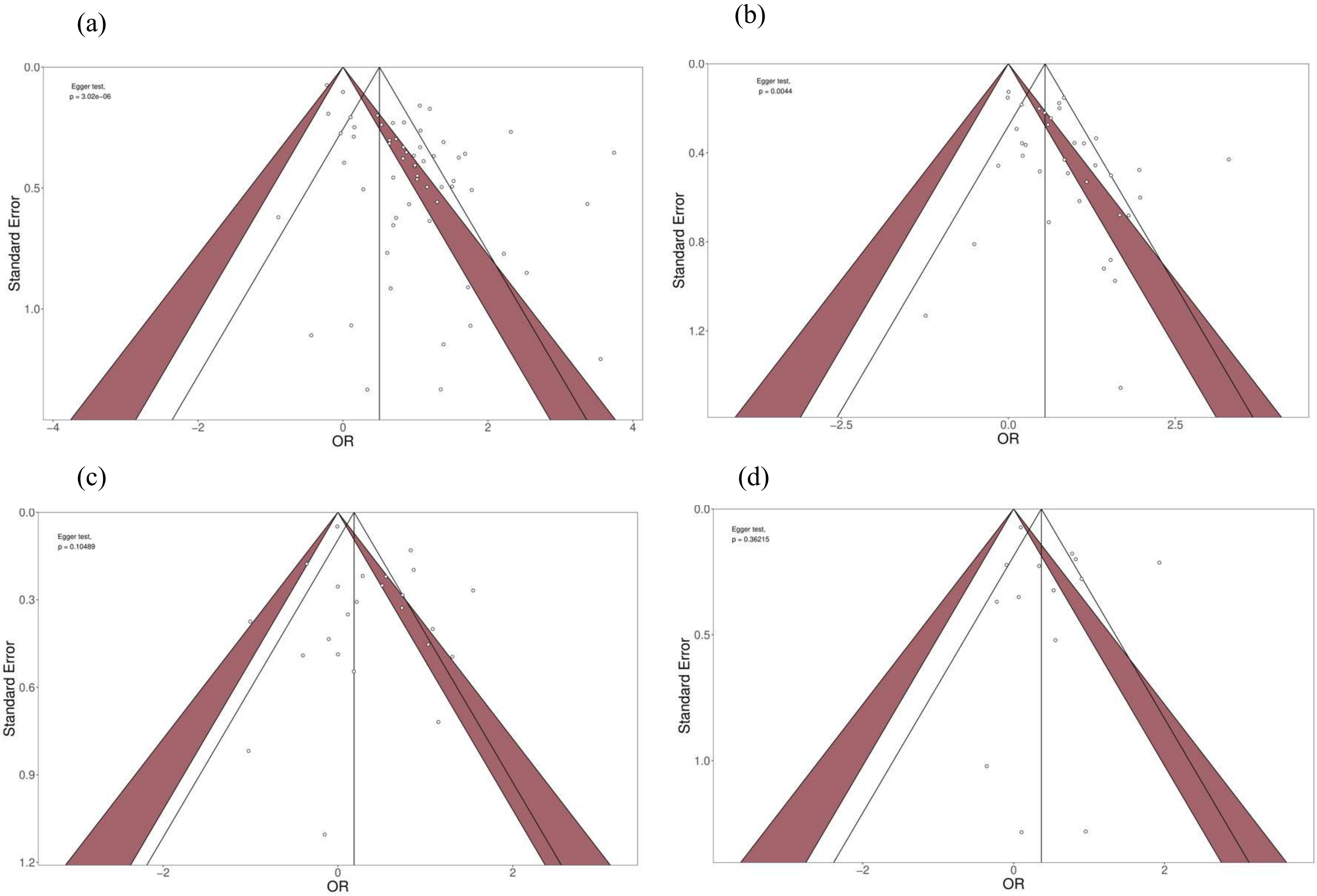

***Figure S5*: Contour-enhanced funnel plots for measures included in the: (a) transfusion, (b) surgery, (c) dental, (d) injection groups.**
These plots show points of individual odds ratios collected in selected studies against associated standard errors. Red-coloured region corresponds to area of statistical significance ($p<0.05$). The solid vertical line represents the overall effect associated with the random-effect model, and the solid triangle represents the associated 95% CI.

**Comments regarding funnel plots**

Overall funnel plot (Fig. S4) suggests potential publication bias as it is asymmetric. This was supported by the Egger test ($p < 0.0001$). Contour-enhanced funnel plots were also computed for 4 procedures groups as they were containing more than 10 measures[1] (Fig. S5): surgery, transfusion, dental, and injection. The Egger test detected asymmetry for measures associated with blood transfusion ($p < 0.0001$) and surgery ($p = 0.0044$). Further inspection of these two funnel plots showed that the asymmetry could be due to potential publication bias. In fact, areas where “missing” studies are expected to restore symmetry do not fall in the region of statistical significance.

(a)

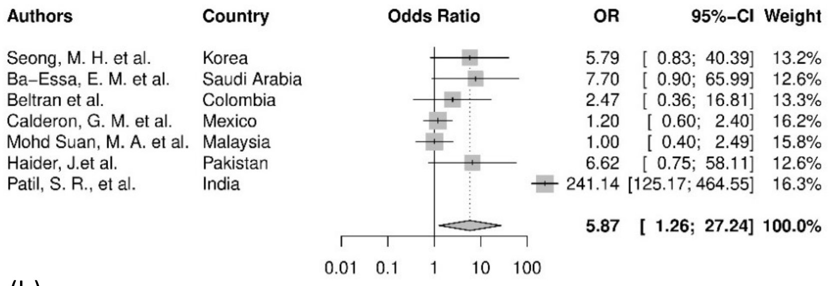

(b)

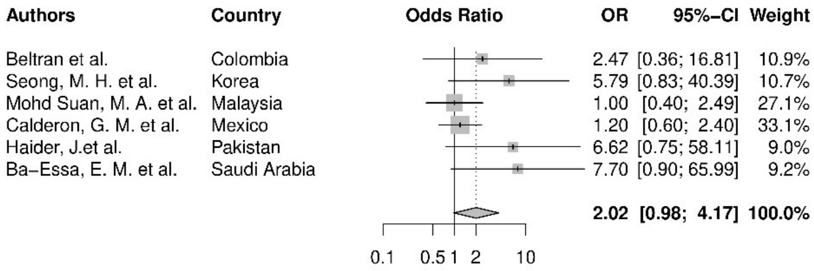

***Figure S6*: Forest plot for the haemodialysis group:** (a) considering the outlier, (b) without the outlier

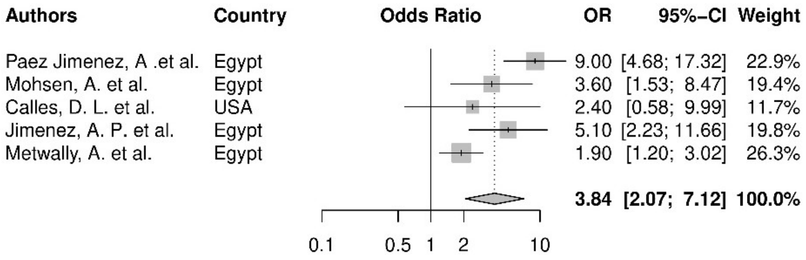

***Figure S7*: Forest plot for the wound care group**

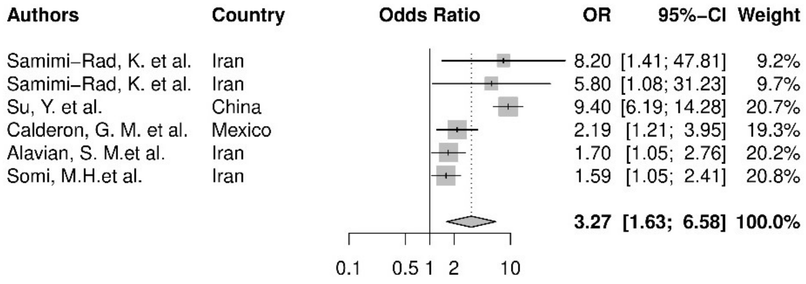

***Figure S8*: Forest plot for the transplantation group**

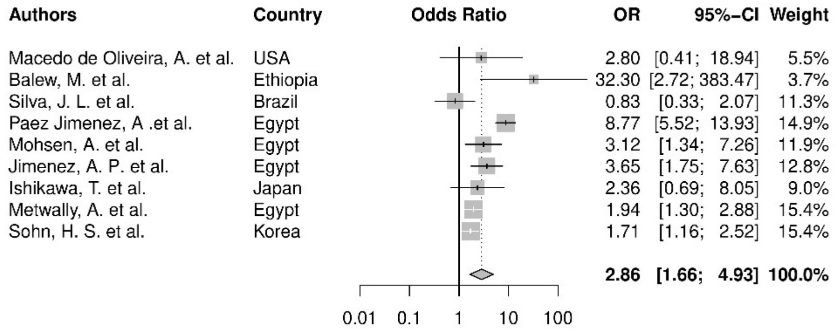

***Figure S9*: Forest plot for the IV/Catheter group**

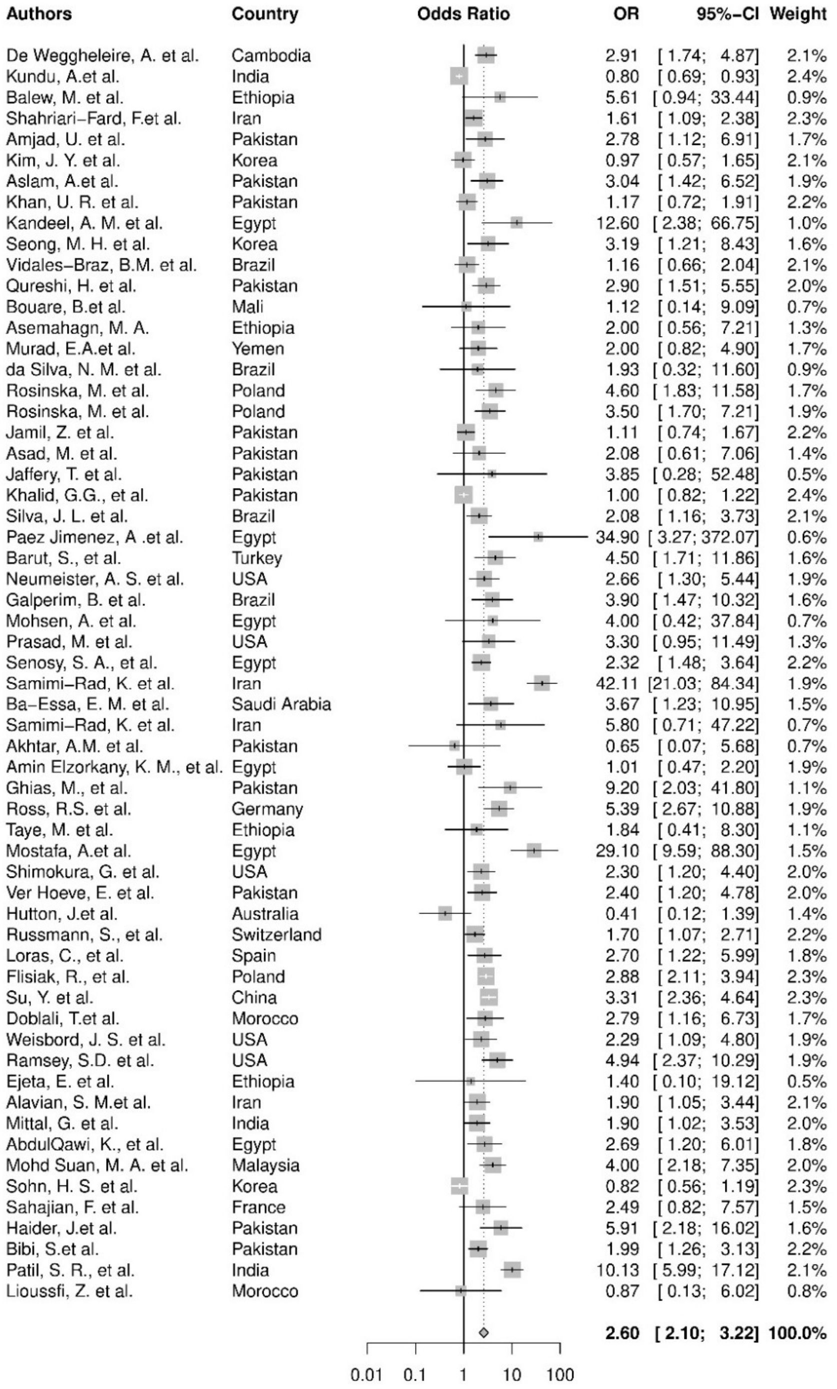

***Figure S10*: Forest plot for the blood transfusion group**

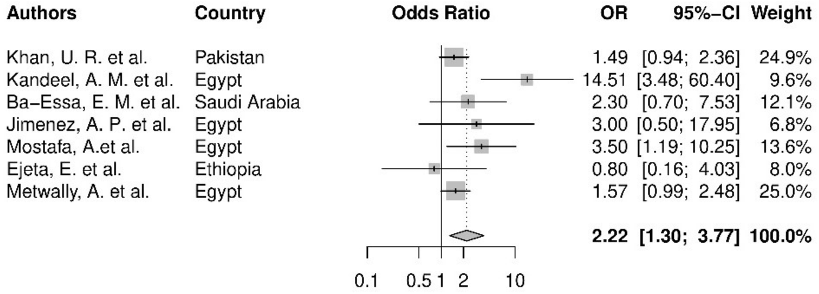

*Figure S11*: **Forest plot for the other procedures group**

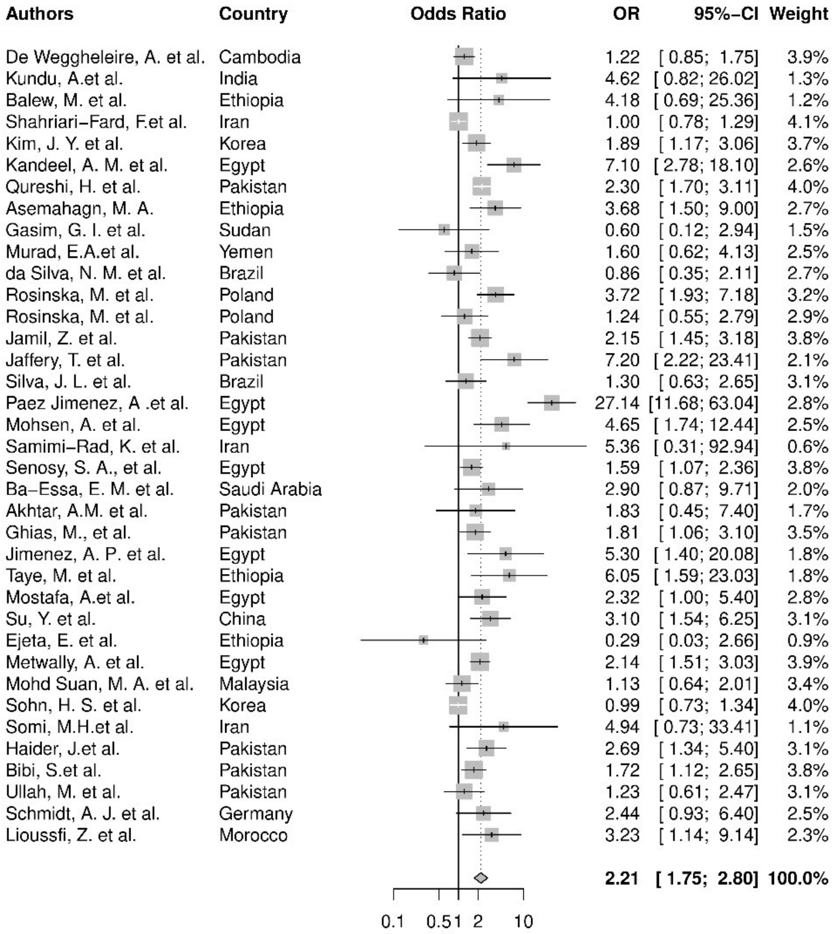

*Figure S12*: **Forest plot for the surgery group**

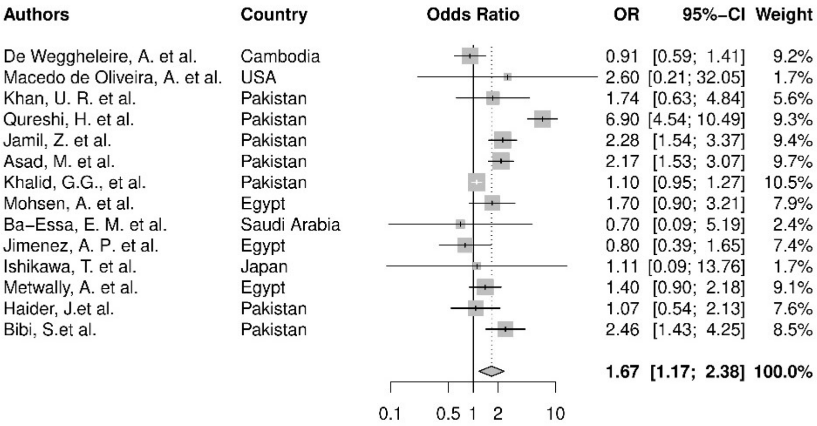

***Figure S13*: Forest plot for the injection group**

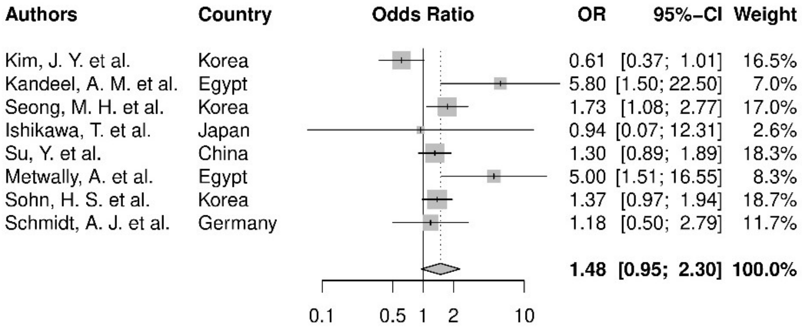

***Figure S14*: Forest plot for the endoscopy group**

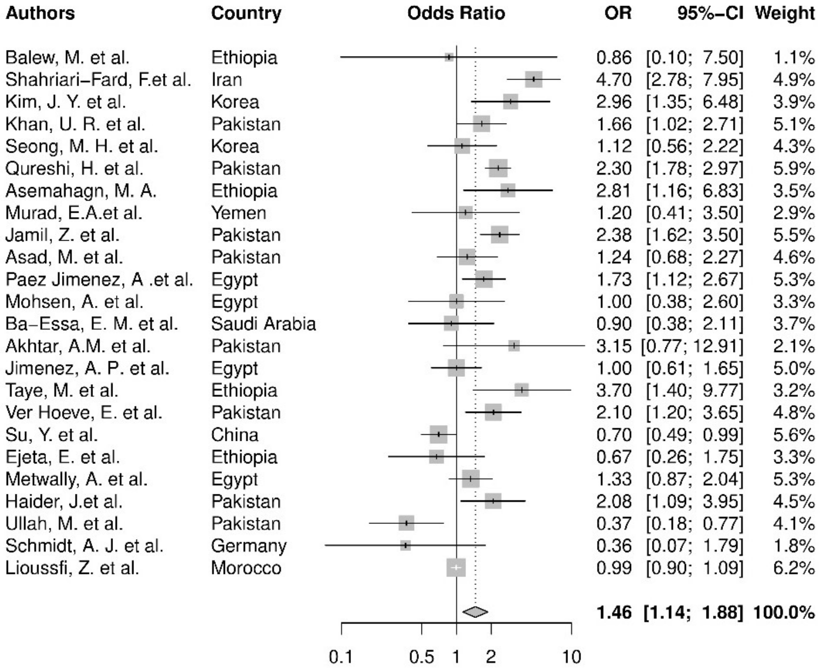

***Figure S15*: Forest plot for the dental procedure group**

**Comments regarding forest plots**

Forest plot are presented without any information on size of HCV positive and HCV negative groups because some measures of association within a group could be estimates provided by a fixed effect model performed when procedure inside same groups were pooled (as described in the Methods section). Each of the measures and associated group size are available in Table S6.

***Table S1*: Search queries used in Scopus, PubMed and Web of Science**

| Database | Full search terms |
|---|---|
| **Scopus** | (TITLE (*"hepatitis" OR* hcv) AND TITLE-ABS-KEY (*"risk factor" OR "odds-ratio" OR "relative risk" OR* "prevalence ratio*"*) AND TITLE-ABS-KEY (hospital* OR nosocomial OR iatrogenic OR healthcare OR practitioner OR clinic OR clinics OR units OR *"care providers")* AND TITLE-ABS-KEY (procedure* OR injection* OR transfusion OR suture OR perfusion OR surgery) AND NOT TITLE-ABS-KEY (pediatric)) AND PUBYEAR > 1999<br>+ filter on article type: remove book chapters, reviews etc.<br>+ filter on language (English or French)<br>+ limit to publications in peer-reviewed journals |
| **PubMed** | ("hepatitis"[Title] OR HCV[Title]) AND ("risk factor"[Title/Abstract] OR "risk factors"[MeSH] OR "prevalence ratio"[Title/Abstract] OR "odds-ratio"[Title/Abstract] OR "relative risk"[Title/Abstract]) AND (hospital*[Title/Abstract] OR nosocomial[Title/Abstract] OR iatrogenic[Title/Abstract] OR healthcare[Title/Abstract] OR practitioner[Title/Abstract] OR clinic[Title/Abstract] OR clinics[Title/Abstract] OR units[Title/Abstract] OR "care providers"[Title/Abstract]) AND (procedure*[Title/Abstract] OR injection*[Title/Abstract] OR transfusion[Title/Abstract] OR suture[Title/Abstract] OR perfusion[Title/Abstract] OR surgery[Title/Abstract]) not(paediatric) NOT(review[PT]) AND ("2000"[Date - Publication] : "2020"[Date - Publication]) |
| **Web of Science** | **TITLE:** ("hepatitis" OR hcv) AND **TOPIC:** ("risk factor" OR "odds-ratio" OR "relative risk"OR "prevalence ratio") AND **TOPIC:** (hospital* OR nosocomial OR iatrogenic OR healthcare OR practitioner OR clinic OR clinics OR units OR «care providers") AND **TOPIC:** (procedure* OR injection* OR transfusion OR suture OR perfusion OR surgery) NOT **TOPIC:** (paediatric)<br>Time span: 2000-2020 |

*Table S2:* **Summary of the bias analysis for each of the selected study for the 9 criteria.**

| Authors | Year | C1 | C2 | C3 | C4 | C5 | C6 | C7 | C8 | C9 |
|---|---|---|---|---|---|---|---|---|---|---|
| Weisbord, J. S. et al. | 2003 | L | PL | PH | L | H | PL | PL | PL | L |
| Alavian, S. M.et al. | 2003 | L | PL | PH | L | H | PL | PL | PL | L |
| Macedo de Oliveira, A. et al. | 2005 | L | PL | L | L | PL | PL | PL | PL | L |
| Jaffery, T. et al. | 2005 | L | PL | PL | L | PH | PL | PL | PL | L |
| Beltran et al. | 2005 | L | PL | PH | L | L | PL | PL | PL | L |
| Ishikawa, T. et al. | 2005 | L | PL | PH | L | PH | PL | PL | PL | L |
| Neumeister, A. S. et al. | 2007 | L | PL | PH | L | H | PL | PL | L | L |
| Samimi-Rad, K. et al. | 2007 | L | PL | PH | L | PL | PL | PL | PL | L |
| Russmann, S., et al. | 2007 | L | PL | PH | L | PL | PL | PL | PL | L |
| Sahajian, F. et al. | 2007 | L | PL | PH | L | PL | PL | PL | PL | L |
| Khan, U. R. et al. | 2008 | L | PL | PH | L | H | PL | PL | PL | L |
| Samimi-Rad, K. et al. | 2008 | L | PL | H | L | PH | PL | PL | PL | L |
| Qureshi, H. et al. | 2009 | L | PL | PH | L | PL | PL | PL | PL | L |
| Ghias, M., et al. | 2009 | L | PL | PH | L | PL | PL | PL | PL | L |
| Jimenez, A. P. et al. | 2009 | L | PL | PH | L | PH | PL | PL | L | L |
| Ross, R.S. et al. | 2009 | L | PL | PH | L | H | PL | PL | L | L |
| Loras, C., et al. | 2009 | L | PL | PH | L | PH | PL | PL | PL | L |
| Calderon, G. M. et al. | 2009 | L | PL | PH | L | L | PL | PL | PL | L |
| Paez Jimenez, A .et al. | 2010 | L | PL | PH | L | PL | PL | PL | L | L |
| Galperim, B. et al. | 2010 | L | PL | PH | L | PL | PL | PL | PL | L |
| AbdulQawi, K., et al. | 2010 | L | PL | PH | L | PH | PL | PL | L | L |
| Silva, J. L. et al. | 2011 | L | PL | PH | L | PH | PL | PL | L | L |
| Barut, S., et al. | 2011 | L | PL | PH | L | PL | PL | PL | PL | L |
| Shimokura, G. et al. | 2011 | L | PL | PH | L | PL | PL | PL | PL | L |
| Flisiak, R., et al. | 2011 | L | PL | PH | L | L | PL | PL | PL | L |
| Schmidt, A. J. et al. | 2011 | L | PL | PH | L | PL | PL | PL | L | L |
| Kim, J. Y. et al. | 2012 | L | PL | PH | L | PL | PL | PL | PL | L |
| Kandeel, A. M. et al. | 2012 | L | PL | PH | L | PH | PL | PL | L | L |
| Gasim, G. I. et al. | 2012 | L | PL | PH | L | L | PL | PL | PL | L |
| Samimi-Rad, K. et al. | 2012 | L | PL | PH | L | PH | PL | PL | PL | L |
| Seong, M. H. et al. | 2013 | L | PL | PH | L | PL | PL | PL | L | L |
| Bouare, B.et al. | 2013 | L | PL | PH | L | H | PL | PL | PL | L |
| Murad, E.A.et al. | 2013 | L | PL | PH | L | PL | PL | PL | PL | L |
| da Silva, N. M. et al. | 2013 | L | PL | PH | L | L | PL | PL | L | L |
| Ver Hoeve, E. et al. | 2013 | L | PL | PH | L | PL | PL | PL | L | L |
| Su, Y. et al. | 2013 | L | PL | PH | L | PL | PL | PL | PL | L |
| Mittal, G. et al. | 2013 | L | PL | PH | L | H | PL | PL | PL | L |
| Bibi, S.et al. | 2013 | L | PL | PH | L | H | PL | PL | PL | L |
| Balew, M. et al. | 2014 | L | PL | PH | L | PH | PL | PL | PL | L |
| Akhtar, A.M. et al. | 2014 | L | PL | PH | L | H | PL | PL | PL | L |
| Doblali, T.et al. | 2014 | L | PL | PH | L | PL | PL | PL | PL | L |

| | | | | | | | | | | |
|---|---|---|---|---|---|---|---|---|---|---|
| Metwally, A. et al. | 2014 | L | PL | PH | L | H | PL | PL | L | L |
| Somi, M.H.et al. | 2014 | L | PL | L | L | H | PL | PL | L | L |
| Lioussfi, Z. et al. | 2014 | L | PL | H | L | PH | PL | PL | PL | L |
| Vidales-Braz, B.M. et al. | 2015 | L | PL | PH | L | PH | PL | PL | PL | L |
| Asad, M. et al. | 2015 | L | PL | PH | L | H | PL | PL | PL | L |
| Mohsen, A. et al. | 2015 | L | PL | PH | L | PL | PL | PL | PL | L |
| Aslam, A.et al. | 2016 | L | PL | PH | L | H | PL | PL | PL | L |
| Senosy, S. A., et al. | 2016 | L | PL | PH | L | PL | PL | PL | PL | L |
| Ba-Essa, E. M. et al. | 2016 | L | PL | PH | L | PH | PL | PL | L | L |
| Sohn, H. S. et al. | 2016 | L | PL | PH | L | PL | PL | PL | L | L |
| Ullah, M. et al. | 2016 | L | PL | PH | L | H | PL | PL | PL | PL |
| De Weggheleire, A. et al. | 2017 | L | PL | PH | L | H | PL | PL | L | L |
| Kundu, A.et al. | 2017 | L | PL | PH | L | H | PL | PL | PL | L |
| Amin Elzorkany, K. M., et al. | 2017 | L | PL | PH | L | PH | PL | PL | PL | L |
| Calles, D. L. et al. | 2017 | L | PL | L | L | L | PL | PL | PL | L |
| Haider, J.et al. | 2017 | L | PL | PH | L | H | PL | PL | L | L |
| Patil, S. R., et al. | 2017 | L | PL | H | L | PL | PL | PL | PL | L |
| Rosinska, M. et al. | 2017 | L | PL | PH | L | L | L | PL | L | L |
| Shahriari-Fard, F.et al. | 2018 | L | PL | PH | PL | H | L | PL | PL | L |
| Khalid, G.G., et al. | 2019 | L | PL | PH | L | PL | L | PL | L | L |
| Taye, M. et al. | 2019 | L | PL | PH | L | PL | PL | PL | L | L |
| Hutton, J.et al. | 2019 | L | PL | PH | L | H | PL | PL | L | L |
| Ramsey, S.D. et al. | 2019 | L | PL | PH | L | H | PL | PL | PL | L |
| Ejeta, E. et al. | 2019 | L | PL | PH | L | PL | PL | PL | L | L |
| Mohd Suan, M. A. et al. | 2019 | L | PL | PH | L | PL | PL | PL | L | L |
| Amjad, U. et al. | 2020 | L | PL | PH | L | H | PL | PL | PL | L |
| Asemahagn, M. A. | 2020 | L | PL | PH | L | PH | PL | PL | L | L |
| Jamil, Z. et al. | 2020 | L | PL | PH | L | H | PL | PL | PL | L |
| Prasad, M. et al. | 2020 | L | PL | PH | L | PL | PL | PL | PL | L |
| Mostafa, A.et al. | 2020 | L | PL | L | L | PH | PL | PL | L | L |

PL: “Probably Low bias”, L: “Low bias”, PH: “Probably High bias”, H: “High bias”

***Table S3:*** **Summary of per-procedure meta-regressions results accounting for the effect of prevalence**

| **Procedure** | **Intercept[se]** | **Prevalence[se]** | **p-value intercept** | **p-value prevalence** |
|---|---|---|---|---|
| haemodialysis | 0·51[0·64] | 0·12[0·24] | 0·43 | 0·63 |
| transplantation | 0·83[0·62] | 0·50[0·70] | 0·18 | 0·47 |
| wound care | 0·80[1·10] | 0·06[0·12] | 0·47 | 0·60 |
| IV/Catheter | 0·54[0·42] | 0·08[0·06] | 0·2 | 0·14 |
| blood transfusion | **0·90[0·16]***** | 0·02[0·03] | <10e-3 | 0·65 |
| endoscopy | 0·03[0·19] | **0·16[0·06]**** | 0·89 | 0·004 |
| surgery | **0·44[0·18]*** | **0·08[0·03]**** | 0·01 | 0·01 |
| dental procedures | 0·38[0·23] | -0·001[0·032] | 0·1 | 0·97 |
| injection | 0·35[0·55] | 0·02[0·08] | 0·52 | 0·76 |
| other procedures | 0·10[0·77] | 0·10[0·09] | 0·89 | 0·28 |

* : 0·01<p<0·05 ; ** : 0·001< p < 0·01 ; *** : p < 0·001

***Table S4:*** **Summary of per-procedure meta-regressions results accounting for the effect of HAQ**

| **Procedure** | **Intercept[se]** | **HAQ[se]** | **p-value intercept** | **p-value prevalence** |
|---|---|---|---|---|
| haemodialysis | -0·41[2·43] | 0·02[0·04] | 0·87 | 0·64 |
| transplantation | -5·10[6·6] | 0·09[0·09] | 0·44 | 0·34 |
| wound care | 3·02[3·20] | -0·03[0·05] | 0·35 | 0·60 |
| IV/Catheter | 3·15[1·56] | -0·03[0·02] | 0·04 | 0·17 |
| blood transfusion | 0·82[0·43] | 0·002[0·006] | 0·06 | 0·75 |
| endoscopy | **4·53[1·75]**** | **-0·05[0·02]*** | 0·01 | 0·02 |
| surgery | 1·06[0·51] | -0·004[0·008] | 0·04 | 0·60 |
| dental procedures | 0·80[0·51] | -0·007[0·008] | 0·12 | 0·40 |
| injection | 1·55[0·82] | -0·02[0·02] | 0·06 | 0·20 |
| other procedures | -0·63[1·46] | 0·02[0·02] | 0·67 | 0·31 |

* : 0·01<p<0·05 ; ** : 0·001< p < 0·01 ; *** : p < 0·001

***Table S5:*** **Summary of overall meta-regressions results for: i) Model 1 (HAQ + Prevalence), ii) Model 2 (HAQ only) and iii) Model 3 (Prevalence only)**

| | Intercept | Prevalence | HAQ | p-value intercept | p-value prevalence | p-value HAQ |
|---|---|---|---|---|---|---|
| All proc· model1 | 0·29[0·30] | **0·05[0·02]*** | 0·005[0·004] | 0·33 | 0·003 | 0·25 |
| All proc· model2 | **0·91[0·23]***** | - | -0·002[0·004] | <10e-3 | - | 0·66 |
| All proc· model3 | **0·62[0·08]***** | **0·04[0·01]*** | - | <10e-3 | 0·005 | - |

* : 0·01<p<0·05 ; ** : 0·001< p < 0·01 ; *** : p < 0·001

*Table S6 :* **Summary table of reported measures within selected studies.**

| **Authors** | **Country** | **Procedure** | **Reported OR/PR/RR** | **CI 95% lower bound** | **CI 95% upper bound** | **Exposed positive** | **Total positive** | **Exposed negative** | **Total negative** | **OR type** |
|---|---|---|---|---|---|---|---|---|---|---|
| De Weggheleire, A. et al. | Cambodia | surgery | 1·22 | 0·85 | 1·75 | 44 | 735 | 113 | 2279 | COR |
| De Weggheleire, A. et al. | Cambodia | injection | 0·91 | 0·59 | 1·41 | 26 | 536 | 131 | 2478 | COR |
| De Weggheleire, A. et al. | Cambodia | transfusion | 2·91 | 1·74 | 4·87 | 36 | 301 | 121 | 2699 | AOR |
| Macedo de Oliveira, A. et al. | USA | central venous catheter | 2·8 | 0·4 | 18·3 | 47 | 56 | 13 | 56 | AOR |
| Macedo de Oliveira, A. et al. | USA | subcutaneous injection | 2·6 | 0·2 | 30·4 | 21 | 56 | 6 | 56 | AOR |
| Kundu, A.et al. | India | surgery | 4·625 | 0·822 | 26·028 | 3 | 9 | 4 | 41 | COR |
| Kundu, A.et al. | India | transfusion | 0·8 | 0·691 | 0·926 | 0 | 9 | 5 | 41 | COR |
| Balew, M. et al. | Ethiopia | surgery | 4·18 | 0·69 | 25·4 | 3 | 5 | 103 | 390 | COR |
| Balew, M. et al. | Ethiopia | catheter | 32·3 | 2·7 | 380·6 | 1 | 5 | 3 | 390 | COR |
| Balew, M. et al. | Ethiopia | dental procedure | 0·86 | 0·1 | 7·6 | 1 | 5 | 88 | 390 | COR |
| Balew, M. et al. | Ethiopia | transfusion | 5·61 | 1·03 | 36·59 | 2 | 5 | 35 | 390 | AOR |
| Shahriari-Fard, F.et al. | Iran | transfusion | 1·61 | 1·09 | 2·39 | 68 | 493 | 47 | 521 | COR |
| Shahriari-Fard, F.et al. | Iran | dental procedure | 4·7 | 2·78 | 7·95 | 476 | 494 | 461 | 543 | COR |
| Shahriari-Fard, F.et al. | Iran | surgery | 1·004 | 0·78 | 1·28 | 226 | 500 | 239 | 530 | COR |
| Amjad, U. et al. | Pakistan | transfusion | 2·78 | 1·118 | 6·902 | 31 | 40 | 31 | 56 | COR |
| Kim, J. Y. et al. | Korea | endoscopy | 0·61 | 0·37 | 1·02 | 579 | 680 | 186 | 206 | COR |
| Kim, J. Y. et al. | Korea | dental care | 2·96 | 1·35 | 6·48 | 631 | 797 | 45 | 85 | AOR |
| Kim, J. Y. et al. | Korea | transfusion after1992 | 0·53 | 0·27 | 1·02 | 78 | 553 | 36 | 195 | AOR |
| Kim, J. Y. et al. | Korea | transfusion before 1992 | 2·99 | 1·2 | 7·41 | 124 | 599 | 11 | 170 | AOR |
| Kim, J. Y. et al. | Korea | surgery | 1·89 | 1·17 | 3·06 | 339 | 709 | 63 | 199 | AOR |
| Aslam, A.et al. | Pakistan | transfusion | 3·04 | 1·43 | 6·57 | 31 | 290 | 11 | 290 | COR |
| Khan, U. R. et al. | Pakistan | abortion | 1·49 | 0·94 | 2·35 | 49 | 119 | 76 | 238 | COR |
| Khan, U. R. et al. | Pakistan | injection | 1·74 | 0·63 | 4·87 | 114 | 335 | 5 | 22 | COR |
| Khan, U. R. et al. | Pakistan | dental procedure | 1·66 | 1·01 | 2·7 | 39 | 93 | 80 | 264 | COR |
| Khan, U. R. et al. | Pakistan | transfusion | 1·17 | 0·72 | 1·91 | 36 | 100 | 83 | 257 | COR |
| Kandeel, A. M. et al. | Egypt | surgery | 7·1 | 2·8 | 18·2 | 16 | 86 | 9 | 287 | COR |
| Kandeel, A. M. et al. | Egypt | endoscopy | 5·8 | 2·1 | 31·6 | 5 | 86 | 3 | 287 | COR |
| Kandeel, A. M. et al. | Egypt | urinary catheter | 6·9 | 1·1 | 55·6 | 4 | 86 | 2 | 287 | COR |
| Kandeel, A. M. et al. | Egypt | transfusion | 12·6 | 3·2 | 89·8 | 7 | 86 | 2 | 287 | COR |
| Kandeel, A. M. et al. | Egypt | abscess drainage | 33·4 | 4·2 | 267·9 | 24 | 86 | 11 | 287 | AOR |
| Seong, M. H. et al. | Korea | haemodialysis | 5·79 | 0·83 | 40·4 | 9 | 1140 | 0 | 526 | AOR |
| Seong, M. H. et al. | Korea | dental care | 1·12 | 0·56 | 2·21 | 1069 | 1145 | 458 | 527 | AOR |
| Seong, M. H. et al. | Korea | endoscopy | 1·73 | 1·08 | 2·76 | 978 | 1148 | 418 | 525 | AOR |
| Seong, M. H. et al. | Korea | transfusion before 1995 | 3·19 | 1·21 | 8·45 | 205 | 1108 | 7 | 525 | AOR |

| | | | | | | | | | | |
|---|---|---|---|---|---|---|---|---|---|---|
| Vidales-Braz, B.M. et al. | Brazil | transfusion | 1·16 | 0·65 | 2·01 | 9 | 11 | 37 | 47 | COR |
| Qureshi, H. et al. | Pakistan | injection | 6·9 | 4·5 | 10·4 | 691 | 723 | 548 | 723 | COR |
| Qureshi, H. et al. | Pakistan | transfusion | 2·9 | 1·5 | 5·5 | 42 | 723 | 15 | 723 | COR |
| Qureshi, H. et al. | Pakistan | dental procedure | 2·3 | 1·8 | 3 | 277 | 723 | 151 | 723 | COR |
| Qureshi, H. et al. | Pakistan | surgery | 2·3 | 1·7 | 3·1 | 164 | 723 | 82 | 723 | COR |
| Bouare, B.et al. | Mali | transfusion | 1·12 | 0·14 | 9·22 | 1 | 13 | 15 | 217 | COR |
| Asemahagn, M. A. | Ethiopia | transfusion | 2 | 0·57 | 7·4 | 3 | 18 | 36 | 404 | COR |
| Asemahagn, M. A. | Ethiopia | tooth extraction | 2·81 | 1·11 | 6·56 | 6 | 18 | 56 | 404 | AOR |
| Asemahagn, M. A. | Ethiopia | surgery | 3·68 | 1·64 | 9·82 | 5 | 18 | 23 | 404 | AOR |
| Gasim, G. I. et al. | Sudan | surgery | 0·6 | 0·1 | 2·4 | NA | 30 | NA | 323 | AOR |
| Murad, E.A.et al. | Yemen | transfusion | 2 | 0·8 | 4·8 | 14 | 34 | 70 | 362 | AOR |
| Murad, E.A.et al. | Yemen | surgery | 1·6 | 0·6 | 4 | 16 | 34 | 91 | 368 | AOR |
| Murad, E.A.et al. | Yemen | dental care | 1·2 | 0·4 | 3·4 | 28 | 34 | 279 | 366 | AOR |
| da Silva, N. M. et al. | Brazil | surgery | 0·86 | 0·35 | 2·11 | 33 | 37 | 110 | 122 | AOR |
| da Silva, N. M. et al. | Brazil | transfusion | 1·93 | 0·32 | 11·56 | 36 | 37 | 114 | 122 | AOR |
| Rosinska, M. et al. | Poland | transfusion before 1992 | 4·6 | 1·83 | 11·59 | NA | 51 | NA | 7529 | AOR |
| Rosinska, M. et al. | Poland | biopsy | 3·72 | 1·93 | 7·19 | NA | 51 | NA | 7529 | AOR |
| Rosinska, M. et al. | Poland | transfusion before 1992 | 3·5 | 1·7 | 7·21 | NA | 51 | NA | 14244 | AOR |
| Rosinska, M. et al. | Poland | caesarean section | 1·24 | 0·55 | 2·78 | NA | 51 | NA | 14244 | AOR |
| Jamil, Z. et al. | Pakistan | transfusion | 1·11 | 0·74 | 1·67 | 156 | 525 | 41 | 149 | COR |
| Jamil, Z. et al. | Pakistan | dental procedure | 2·38 | 1·62 | 3·51 | 271 | 525 | 46 | 149 | COR |
| Jamil, Z. et al. | Pakistan | injection | 2·28 | 1·54 | 3·37 | 261 | 525 | 45 | 149 | COR |
| Jamil, Z. et al. | Pakistan | surgery | 2·15 | 1·45 | 3·18 | 249 | 525 | 44 | 149 | COR |
| Asad, M. et al. | Pakistan | injection | 2·17 | 1·53 | 3·07 | 112 | 112 | 0 | 233 | COR |
| Asad, M. et al. | Pakistan | dental procedure | 1·24 | 0·68 | 2·27 | 15 | 112 | NA | 233 | COR |
| Asad, M. et al. | Pakistan | transfusion | 2·08 | 0·61 | 7·03 | 5 | 112 | NA | 233 | COR |
| Jaffery, T. et al. | Pakistan | surgery | 7·204 | 2·2 | 23·23 | 12 | 31 | 7 | 62 | AOR |
| Jaffery, T. et al. | Pakistan | transfusion | 3·85 | 0·28 | 52·03 | 3 | 31 | 2 | 62 | AOR |
| Khalid, G.G., et al. | Pakistan | transfusion | 1 | 0·8 | 1·2 | 203 | 1847 | 305 | 3145 | AOR |
| Khalid, G.G., et al. | Pakistan | injection | 1·1 | 0·9 | 1·2 | 1141 | 1847 | 1692 | 3145 | AOR |
| Silva, J. L. et al. | Brazil | transfusion | 2·08 | 0·7 | 2·25 | 43 | 69 | 101 | 161 | COR |
| Silva, J. L. et al. | Brazil | sclerotherapy | 0·83 | 0·33 | 2·06 | 8 | 17 | 114 | 213 | COR |
| Silva, J. L. et al. | Brazil | ligation of esophageal varice | 0·35 | 0·05 | 2·53 | 1 | 17 | 34 | 213 | COR |
| Silva, J. L. et al. | Brazil | surgery | 2 | 0·48 | 8·42 | 15 | 17 | 167 | 213 | COR |
| Silva, J. L. et al. | Brazil | splenectomy | 1·44 | 0·58 | 3·58 | 9 | 17 | 96 | 213 | COR |
| Paez Jimenez, A .et al. | Egypt | catheter | 28·4 | 9·4 | 85·7 | 27 | 100 | 39 | 678 | COR |
| Paez Jimenez, A .et al. | Egypt | transfusion | 34·9 | 3·3 | 375·1 | 4 | 100 | 1 | 679 | COR |
| Paez Jimenez, A .et al. | Egypt | caesarean section | 84·2 | 12 | 589·1 | 8 | 100 | 3 | 678 | COR |
| Paez Jimenez, A .et al. | Egypt | IV line | 13·3 | 6·8 | 25·8 | 36 | 100 | 39 | 678 | COR |

| | | | | | | | | | |
|---|---|---|---|---|---|---|---|---|---|
| Paez Jimenez, A .et al. | Egypt | intravenous injection | 2·7 | 1·2 | 5·8 | 13 | 100 | 33 | 678 | COR |
| Paez Jimenez, A .et al. | Egypt | tooth filling | 1·6 | 0·5 | 5·2 | 4 | 100 | 22 | 678 | COR |
| Paez Jimenez, A .et al. | Egypt | wound suture | 9 | 4·7 | 17·4 | 15 | 100 | 6 | 678 | COR |
| Paez Jimenez, A .et al. | Egypt | dental anaesthesia | 1·9 | 1 | 3·6 | 19 | 100 | 51 | 678 | COR |
| Paez Jimenez, A .et al. | Egypt | tooth extraction | 1·6 | 0·8 | 3·1 | 15 | 100 | 55 | 678 | COR |
| Paez Jimenez, A .et al. | Egypt | surgery | 20·9 | 8·2 | 53·2 | 21 | 100 | 10 | 678 | COR |
| Barut, S., et al. | Turkey | transfusion before 1996 | 4·5 | 1·7 | 11·8 | 35 | 189 | 9 | 190 | AOR |
| Neumeister, A. S. et al. | USA | transfusion | 2·7 | 1·041 | 6·971 | 8 | 21 | 38 | 222 | COR |
| Neumeister, A. S. et al. | USA | transfusion before 1992 | 2·6 | 0·8739 | 7·787 | 5 | 21 | 24 | 222 | COR |
| Galperim, B. et al. | Brazil | transfusion | 3·9 | 1·5 | 10·5 | 60 | 68 | 187 | 257 | AOR |
| Mohsen, A. et al. | Egypt | intravenous injection | 3·2 | 1·1 | 10 | 14 | 111 | 5 | 111 | COR |
| Mohsen, A. et al. | Egypt | intramuscular injection | 1·7 | 0·9 | 3·2 | 30 | 111 | 20 | 111 | COR |
| Mohsen, A. et al. | Egypt | dental procedure | 1 | 0·4 | 2·7 | 12 | 111 | 13 | 111 | COR |
| Mohsen, A. et al. | Egypt | catheter | 3 | 0·8 | 11·1 | 9 | 111 | 3 | 111 | COR |
| Mohsen, A. et al. | Egypt | transfusion | 4 | 0·4 | 35·8 | 4 | 111 | 1 | 111 | COR |
| Mohsen, A. et al. | Egypt | caesarean section | 10 | 1·3 | 78·1 | 10 | 111 | 1 | 111 | COR |
| Mohsen, A. et al. | Egypt | surgery | 3·7 | 1·2 | 11·3 | 15 | 111 | 4 | 111 | COR |
| Mohsen, A. et al. | Egypt | wound suture | 3·6 | 1·5 | 8·3 | 27 | 111 | 10 | 111 | COR |
| Prasad, M. et al. | USA | transfusion | 3·3 | 0·94 | 11·4 | 13 | 131 | 10 | 251 | AOR |
| Samimi-Rad, K. et al. | Iran | splenectomy | 5·36 | 0·31 | 93·21 | 4 | 32 | 1 | 66 | AOR |
| Senosy, S. A., et al. | Egypt | transfusion | 2·32 | 1·48 | 3·637 | 21 | 37 | 35 | 343 | AOR |
| Senosy, S. A., et al. | Egypt | surgery | 1·586 | 0·998 | 2·204 | 22 | 37 | 165 | 343 | AOR |
| Samimi-Rad, K. et al. | Iran | transfusion | 1·59 | 0·18 | 13·7 | 6 | 7 | 121 | 153 | COR |
| Samimi-Rad, K. et al. | Iran | transfusion before 1996 | 61·3 | 6·9 | 29·9 | 3 | 4 | 14 | 46 | AOR |
| Samimi-Rad, K. et al. | Iran | renal transplantation | 8·2 | 1·4 | 47·6 | 3 | 7 | 7 | 153 | AOR |
| Ba-Essa, E. M. et al. | Saudi Arabia | abortion | 2·3 | 0·7 | 7·5 | 9 | 14 | 314 | 633 | COR |
| Ba-Essa, E. M. et al. | Saudi Arabia | transfusion | 3·67 | 1·23 | 10·95 | 9 | 20 | 176 | 1037 | AOR |
| Ba-Essa, E. M. et al. | Saudi Arabia | injection | 0·7 | 0·1 | 5·5 | 1 | 20 | 70 | 1037 | COR |
| Ba-Essa, E. M. et al. | Saudi Arabia | surgery | 2·9 | 0·9 | 10·1 | 17 | 20 | 683 | 1037 | COR |
| Ba-Essa, E. M. et al. | Saudi Arabia | haemodialysis | 7·7 | 0·9 | 66·1 | 1 | 20 | 7 | 1037 | AOR |
| Ba-Essa, E. M. et al. | Saudi Arabia | dental procedure | 0·9 | 0·4 | 2·2 | 11 | 20 | 599 | 1037 | COR |
| Samimi-Rad, K. et al. | Iran | transfusion | 5·8 | 0·7 | 46·4 | 10 | 11 | 122 | 193 | COR |
| Samimi-Rad, K. et al. | Iran | renal transplantation | 5·8 | 1 | 29 | 3 | 11 | 10 | 193 | AOR |
| Akhtar, A.M. et al. | Pakistan | surgery | 1·833 | 0·454 | 7·391 | 5 | 9 | 30 | 74 | COR |
| Akhtar, A.M. et al. | Pakistan | transfusion | 0·6458 | 0·073 | 5·649 | 1 | 9 | 12 | 74 | COR |
| Akhtar, A.M. et al. | Pakistan | dental procedure | 3·155 | 0·77 | 12·9 | 5 | 26 | 4 | 57 | COR |
| Beltran et al. | Colombia | haemodialysis | 2·47 | 0·41 | 19 | 5 | 45 | 77 | 455 | COR |

| | | | | | | | | | | |
|---|---|---|---|---|---|---|---|---|---|---|
| Amin Elzorkany, K. M., et al. | Egypt | transfusion | 1·015 | 0·467 | 2·204 | 51 | 61 | 415 | 590 | AOR |
| Calles, D. L. et al. | USA | wound care | 2·4 | 0·6 | 10·4 | 25 | 53 | 5 | 39 | AOR |
| Ghias, M., et al. | Pakistan | transfusion | 9·204 | 2·027 | 41·803 | 89 | 281 | 24 | 119 | AOR |
| Ghias, M., et al. | Pakistan | operation | 1·81 | 1·058 | 3·096 | 126 | 281 | 29 | 119 | AOR |
| Jimenez, A. P. et al. | Egypt | intramuscular injection | 0·8 | 0·4 | 1·7 | 13 | 94 | 30 | 188 | COR |
| Jimenez, A. P. et al. | Egypt | abscess drainage | 3 | 0·5 | 17·9 | 3 | 95 | 2 | 188 | COR |
| Jimenez, A. P. et al. | Egypt | dental procedure | 1 | 0·7 | 1·9 | 16 | 94 | 31 | 188 | COR |
| Jimenez, A. P. et al. | Egypt | intravenous injection | 4·3 | 1·3 | 14 | 9 | 94 | 4 | 188 | COR |
| Jimenez, A. P. et al. | Egypt | cannula | 3·3 | 1·3 | 8·5 | 13 | 94 | 9 | 188 | COR |
| Jimenez, A. P. et al. | Egypt | surgery | 5·3 | 1·4 | 20·1 | 8 | 94 | 3 | 188 | COR |
| Jimenez, A. P. et al. | Egypt | wound suture | 5·1 | 2·2 | 11·5 | 21 | 94 | 9 | 188 | COR |
| Ross, R.S. et al. | Germany | transfusion before 1991 | 5·389 | 2·668 | 10·885 | NA | 150 | NA | 150 | COR |
| Taye, M. et al. | Ethiopia | transfusion | 1·84 | 0·41 | 8·34 | 3 | 23 | 22 | 422 | AOR |
| Taye, M. et al. | Ethiopia | surgery | 6·05 | 1·59 | 23·04 | 4 | 23 | 21 | 422 | AOR |
| Taye, M. et al. | Ethiopia | dental care | 3·7 | 1·4 | 9·77 | 8 | 23 | 51 | 422 | AOR |
| Mostafa, A.et al. | Egypt | surgery | 4·4 | 1 | 19·3 | 2 | 22 | 48 | 2155 | COR |
| Mostafa, A.et al. | Egypt | transfusion | 29·1 | 9·6 | 88·4 | 7 | 22 | 28 | 2155 | AOR |
| Mostafa, A.et al. | Egypt | caesarean section | 1·7 | 0·6 | 4·7 | 14 | 22 | 748 | 2155 | AOR |
| Mostafa, A.et al. | Egypt | abortion | 3·5 | 1·2 | 10·3 | 5 | 22 | 197 | 2155 | AOR |
| Ishikawa, T. et al. | Japan | cholangiography | 2·5 | 0·47 | 13·27 | 6 | 14 | 3 | 13 | COR |
| Ishikawa, T. et al. | Japan | intravenous pyelography | 2·5 | 0·24 | 25·68 | 9 | 24 | 0 | 3 | COR |
| Ishikawa, T. et al. | Japan | percutaneaous injection | 1·11 | 0·09 | 13·84 | 9 | 26 | 0 | 1 | COR |
| Ishikawa, T. et al. | Japan | endoscopy | 0·94 | 0·07 | 12 | 1 | 3 | 8 | 23 | COR |
| Ishikawa, T. et al. | Japan | intravenous injection | 1·8 | 0·1 | 31·9 | 0 | 0 | 9 | 26 | COR |
| Shimokura, G. et al. | USA | transfusion before 1992 | 2·3 | 1·2 | 4·4 | 86 | 291 | 507 | 2642 | AOR |
| Ver Hoeve, E. et al. | Pakistan | transfusion | 2·397 | 1·201 | 4·785 | 95 | 145 | 205 | 455 | AOR |
| Ver Hoeve, E. et al. | Pakistan | dental care | 2·095 | 1·201 | 3·655 | 166 | 257 | 134 | 343 | AOR |
| Hutton, J.et al. | Australia | transfusion before 1990 | 0·41 | 0·12 | 1·37 | 3 | 50 | 43 | 318 | COR |
| Russmann, S., et al. | Switzerland | transfusion | 1·7 | 1·1 | 2·8 | 38 | 135 | 945 | 4592 | AOR |
| Loras, C., et al. | Spain | transfusion | 2·7 | 1·2 | 5·9 | NA | 37 | NA | 2039 | AOR |
| Flisiak, R., et al. | Poland | transfusion before 1992 | 2·88 | 2·08 | 3·89 | NA | 354 | NA | 17879 | AOR |
| Su, Y. et al. | China | endoscopy | 1·3 | 0·9 | 1·9 | 46 | 129 | 584 | 1983 | COR |
| Su, Y. et al. | China | dental procedure | 0·7 | 0·5 | 1 | 68 | 129 | 1240 | 2003 | COR |
| Su, Y. et al. | China | surgery | 3·1 | 1·5 | 6·1 | 119 | 129 | 1657 | 1980 | AOR |
| Su, Y. et al. | China | renal transplantation | 9·4 | 6·2 | 14·3 | 42 | 129 | 97 | 1987 | COR |
| Su, Y. et al. | China | transfusion before 1998 | 6·3 | 3·6 | 11·1 | 61 | 83 | 180 | 1258 | AOR |
| Su, Y. et al. | China | transfusion after1998 | 2·3 | 1·5 | 3·5 | 39 | 61 | 657 | 1735 | AOR |
| Doblali, T.et al. | Morocco | transfusion | 2·79 | 1·15 | 6·7 | 22 | 26 | 78 | 115 | COR |

| | | | | | | | | | | |
|---|---|---|---|---|---|---|---|---|---|---|
| Calderon, G. M. et al. | Mexico | haemodialysis | 1·2 | 0·6 | 2·4 | 14 | 41 | 77 | 259 | COR |
| Calderon, G. M. et al. | Mexico | transplantation | 1·6 | 0·9 | 3·8 | 6 | 41 | 25 | 259 | COR |
| Calderon, G. M. et al. | Mexico | transplantation | 4·2 | 1·4 | 11·2 | 6 | 41 | 25 | 259 | AOR |
| Weisbord, J. S. et al. | USA | transfusion | 2·02 | 0·824 | 6·63 | 4 | 32 | 41 | 655 | COR |
| Weisbord, J. S. et al. | USA | transfusion before 1992 | 2·6 | 1·055 | 8·7 | 4 | 32 | 31 | 654 | COR |
| Ramsey, S.D. et al. | USA | blood transfusion 1978 1985 | 4·94 | 2·27 | 9·85 | 7 | 71 | 61 | 2929 | COR |
| Ejeta, E. et al. | Ethiopia | tooth extraction | 0·67 | 0·26 | 1·78 | 6 | 34 | 96 | 387 | AOR |
| Ejeta, E. et al. | Ethiopia | abortion | 0·8 | 0·16 | 4·06 | 2 | 34 | 33 | 387 | AOR |
| Ejeta, E. et al. | Ethiopia | transfusion | 1·4 | 0·1 | 18·66 | 1 | 34 | 7 | 387 | AOR |
| Ejeta, E. et al. | Ethiopia | caesarean section | 0·29 | 0·03 | 2·53 | 1 | 34 | 37 | 387 | AOR |
| Alavian, S. M.et al. | Iran | renal transplantation | 1·7 | 1·1 | 2·9 | 27 | 104 | 105 | 641 | AOR |
| Alavian, S. M.et al. | Iran | transfusion | 1·9 | 1·1 | 3·6 | 96 | 108 | 517 | 642 | COR |
| Metwally, A. et al. | Egypt | surgery | 1·378 | 0·9 | 2·1 | 292 | 540 | 47 | 102 | COR |
| Metwally, A. et al. | Egypt | dental procedure | 1·328 | 0·85 | 2 | 378 | 540 | 65 | 102 | COR |
| Metwally, A. et al. | Egypt | catheter | 1·31 | 0·58 | 3·07 | 54 | 540 | 8 | 102 | COR |
| Metwally, A. et al. | Egypt | laparoscopy | 2·288 | 1·07 | 4·87 | 88 | 540 | 8 | 102 | COR |
| Metwally, A. et al. | Egypt | cannula | 2·273 | 1·41 | 3·64 | 243 | 540 | 27 | 102 | COR |
| Metwally, A. et al. | Egypt | wound suture | 1·9 | 1·19 | 3 | 226 | 540 | 28 | 102 | COR |
| Metwally, A. et al. | Egypt | abscess drainage | 1·2 | 0·72 | 2·21 | 109 | 540 | 17 | 102 | COR |
| Metwally, A. et al. | Egypt | endoscopy | 5 | 1·5 | 16·44 | 72 | 540 | 3 | 103 | COR |
| Metwally, A. et al. | Egypt | sclerotherapy | 1·4 | 0·32 | 6·34 | 15 | 540 | 2 | 102 | COR |
| Metwally, A. et al. | Egypt | injection vaccination | 1·4 | 0·9 | 2·19 | 230 | 540 | 35 | 102 | COR |
| Metwally, A. et al. | Egypt | organ biopsy | 23·6 | 8·56 | 65 | 265 | 540 | 4 | 102 | COR |
| Metwally, A. et al. | Egypt | tapping ascites | 6·36 | 0·86 | 47 | 32 | 540 | 1 | 102 | COR |
| Metwally, A. et al. | Egypt | hemorrhoids treatment | 3·5 | 1 | 11·4 | 52 | 540 | 3 | 102 | COR |
| Metwally, A. et al. | Egypt | electromyogram | 1·46 | 0·43 | 4·98 | 23 | 540 | 3 | 102 | COR |
| Mittal, G. et al. | India | transfusion | 1·9 | 0·69 | 2·38 | 26 | 36 | 42 | 82 | COR |
| AbdulQawi, K., et al. | Egypt | transfusion | 2·69 | 1·2 | 6 | 12 | 83 | 45 | 1141 | AOR |
| Mohd Suan, M. A. et al. | Malaysia | transfusion before 1992 | 4 | 2·24 | 7·56 | 51 | 66 | 204 | 444 | COR |
| Mohd Suan, M. A. et al. | Malaysia | surgery | 1·13 | 0·64 | 2·02 | 28 | 53 | 227 | 457 | COR |
| Mohd Suan, M. A. et al. | Malaysia | haemodialysis | 1 | 0·4 | 2·48 | 10 | 20 | 245 | 490 | COR |
| Sohn, H. S. et al. | Korea | gastroscopy | 1·15 | 0·55 | 2·43 | 216 | 234 | 214 | 234 | COR |
| Sohn, H. S. et al. | Korea | colonoscopy | 1·44 | 0·98 | 2·13 | 135 | 234 | 116 | 234 | COR |
| Sohn, H. S. et al. | Korea | transfusion | 0·76 | 0·51 | 1·12 | 64 | 234 | 78 | 234 | COR |
| Sohn, H. S. et al. | Korea | surgery | 1·15 | 0·78 | 1·7 | 152 | 234 | 144 | 234 | COR |
| Sohn, H. S. et al. | Korea | bloody operation | 0·8 | 0·5 | 1·27 | 40 | 234 | 48 | 234 | COR |
| Sohn, H. S. et al. | Korea | phlebotomy | 1·71 | 1·16 | 2·51 | 106 | 234 | 78 | 234 | COR |
| Sohn, H. S. et al. | Korea | transfusion before 1990 | 2 | 0·5 | 8 | 24 | 234 | 16 | 234 | COR |

| | | | | | | | | | | |
|---|---|---|---|---|---|---|---|---|---|---|
| Somi, M.H.et al. | Iran | renal transplantation | 1·59 | 1·05 | 2·41 | 8 | 37 | 49 | 418 | COR |
| Somi, M.H.et al. | Iran | surgery | 4·94 | 0·73 | 33·4 | 17 | 37 | 317 | 418 | COR |
| Sahajian, F. et al. | France | transfusion before 1992 | 2·49 | 0·73 | 6·75 | 5 | 44 | 44 | 897 | COR |
| Haider, J.et al. | Pakistan | injection | 1·07 | 0·54 | 2·13 | 11 | 46 | 340 | 1494 | COR |
| Haider, J.et al. | Pakistan | transfusion | 5·91 | 2·18 | 16·01 | 5 | 46 | 30 | 1494 | COR |
| Haider, J.et al. | Pakistan | surgery | 2·69 | 1·34 | 5·41 | 11 | 46 | 156 | 1494 | COR |
| Haider, J.et al. | Pakistan | dental procedure | 2·08 | 1·09 | 3·94 | 14 | 46 | 260 | 1494 | COR |
| Haider, J.et al. | Pakistan | haemodialysis | 6·62 | 0·75 | 57·8 | 1 | 46 | 5 | 1494 | COR |
| Bibi, S.et al. | Pakistan | transfusion | 1·99 | 1·26 | 3·12 | 102 | 146 | 106 | 197 | COR |
| Bibi, S.et al. | Pakistan | injection | 2·46 | 1·43 | 4·26 | 124 | 146 | 137 | 197 | COR |
| Bibi, S.et al. | Pakistan | surgery | 1·72 | 1·12 | 2·66 | 85 | 146 | 88 | 197 | COR |
| Ullah, M. et al. | Pakistan | dental procedure | 0·367 | 0·176 | 0·7648 | 31 | 45 | 193 | 225 | COR |
| Ullah, M. et al. | Pakistan | surgery | 1·227 | 0·6093 | 2·472 | 73 | 86 | 151 | 184 | COR |
| Patil, S. R., et al. | India | transfusion | 10·128 | 5·991 | 17·123 | NA | 116 | NA | 25077 | AOR |
| Patil, S. R., et al. | India | haemodialysis | 241·135 | 125 | 463·951 | NA | 116 | NA | 25077 | AOR |
| Schmidt, A. J. et al. | Germany | endoscopy | 1·18 | 0·5 | 2·79 | 13 | 34 | 23 | 67 | COR |
| Schmidt, A. J. et al. | Germany | surgery | 2·44 | 0·93 | 6·4 | 11 | 34 | 11 | 67 | COR |
| Schmidt, A. J. et al. | Germany | dental procedure | 0·36 | 0·07 | 1·73 | 2 | 34 | 10 | 67 | COR |
| Lioussfi, Z. et al. | Morocco | surgery | 3·231 | 1·142 | 9·143 | 30 | 40 | 13 | 27 | COR |
| Lioussfi, Z. et al. | Morocco | dental procedure | 0·993 | 0·903 | 1·092 | 27 | 40 | 22 | 27 | COR |
| Lioussfi, Z. et al. | Morocco | transfusion | 0·868 | 0·125 | 6·013 | 32 | 40 | 18 | 27 | AOR |

COR : Crude odds ratio ; AOR : Adjusted odds ratio ; NA : Information was not available

***Table S7* : Prevalence and HAQ index scores for each country represented in the meta-analysis**

| Country | Prevalence (%)[2] | HAQ Index score[3] |
|---|---|---|
| Australia | 1·4 | 89·8 |
| Brazil | 0·7 | 64·9 |
| Cambodia | 2·3 | 50·7 |
| China | 1·3 | 74·2 |
| Colombia | 0·9 | 67·8 |
| Egypt | 10 | 61 |
| Ethiopia | 2·7 | 44·2 |
| France | 0·8 | 87·9 |
| Germany | 0·6 | 86·4 |
| India | 0·8 | 44·8 |
| Iran | 0·3 | 71·1 |
| Japan | 1·5 | 89 |
| South Korea | 0·8 | 85·8 |
| Malaysia | 2·5 | 66·6 |
| Mali | 5 | 45·6 |
| Mexico | 1·4 | 62·6 |
| Morocco | 1·2 | 61·3 |
| Pakistan | 6·8 | 43·1 |
| Poland | 0·86 | 79·6 |
| Saudi Arabia | 1·5 | 79·4 |
| Spain | 1·7 | 89·6 |
| Sudan | 1·7 | 50·1 |
| Switzerland | 1·5 | 91·8 |
| Turkey | 1 | 76·2 |
| USA | 1·3 | 81·3 |
| Yemen | 2·2 | 49·6 |
